# Large-Scale Validation Study of an Improved Semi-Autonomous Urine Cytology Assessment Tool: AutoParis-X

**DOI:** 10.1101/2023.03.01.23286639

**Authors:** Joshua J. Levy, Natt Chan, Jonathan D. Marotti, Darcy A. Kerr, Edward J. Gutmann, Ryan E. Glass, Caroline P. Dodge, Arief A. Suriawinata, Brock Christensen, Xiaoying Liu, Louis J. Vaickus

## Abstract

Adopting a computational approach for the assessment of urine cytology specimens has the potential to improve the efficiency, accuracy and reliability of bladder cancer screening, which has heretofore relied on semi-subjective manual assessment methods. As rigorous, quantitative criteria and guidelines have been introduced for improving screening practices, e.g., The Paris System for Reporting Urinary Cytology (TPS), algorithms to emulate semi-autonomous diagnostic decision-making have lagged behind, in part due to the complex and nuanced nature of urine cytology reporting. In this study, we report on a deep learning tool, AutoParis-X, which can facilitate rapid semi-autonomous examination of urine cytology specimens. Through a large-scale retrospective validation study, results indicate that AutoParis-X can accurately determine urothelial cell atypia and aggregate a wide-variety of cell and cluster-related information across a slide to yield an Atypia Burden Score (ABS) that correlates closely with overall specimen atypia, predictive of TPS diagnostic categories. Importantly, this approach accounts for challenges associated with assessment of overlapping cell cluster borders, which improved the ability to predict specimen atypia and accurately estimate the nuclear-to-cytoplasm (NC) ratio for cells in these clusters. We developed an interactive web application that is publicly available and open-source, which features a simple, easy-to-use display for examining urine cytology whole-slide images (WSI) and determining the atypia level of specific cells, flagging the most abnormal cells for pathologist review. The accuracy of AutoParis-X (and other semi-automated digital pathology systems) indicates that these technologies are approaching clinical readiness and necessitates full evaluation of these algorithms via head-to-head clinical trials.

## Introduction

Urothelial carcinoma is highly prevalent (9^th^ most common worldwide) and has the highest recurrence rate among all forms of cancer (74%) ^1, 2^. The treatment and management of urothelial carcinoma requires follow-up urine cytology (UC), expensive, painful chemotherapy, and/or invasive cystoscopy procedures for long periods of time (typically the remainder of the patient’s life), necessitating the development and implementation of less invasive screening and follow up measures ^3^.

The detection and screening for bladder cancer has greatly improved since the earliest recorded evaluation of hematuria was recorded in the papyrus of Kahun, circa 1900 B.C.. In 1550 B.C., it was suggested that hematuria originated from “worms in the belly” ^4^. A causative agent, *S. haematobium*, was identified in 1854 by Theodor Bilharz ^5, 6^. In 1947, Dr. George Papanicolaou, widely considered the father of modern cytopathology, proposed a formal system for evaluation of malignant cells exfoliated from the bladder’s epithelium, which has largely remained intact ^7, 8^. Over the past half-century, efforts to rigorously define quantitative assessment criteria (e.g., nuclear-to-cytoplasm (NC) ratio, chromatin structure, etc.) and improve specimen preparation methods have sought to resolve remaining ambiguity. Yet, traditional cytological approaches are still hampered by inter-rater variability, specimen quality issues, and the tendency towards ‘hedging’ to the atypical category ^9–12^.

In recent years, The Paris System for Reporting Urinary Cytology (TPS), formulated in 2013, published in 2016, and updated in 2022, has emerged as a more quantitative and reproducible reporting system bladder cancer ^13–17^. TPS criteria are applied to assign one of four main ordered categories (negative, atypical urothelial cells, suspicious for high-grade urothelial carcinoma, positive for high-grade urothelial carcinoma) based on the following criteria for a positive diagnosis: (1) at least five malignant urothelial cells (updated to ten in 2022), (2) an NC ratio at or above 0.7, (3) nuclear hyperchromasia, (4) markedly irregular nuclear membrane, and (5) coarse/clumped chromatin ^2^. It is often easier to evaluate specimens that have clear-cut diagnoses, either negative or positive, than those that are atypical or suspicious. Atypical specimens are those that are hedged against a negative diagnosis, while suspicious specimens are those that are hedged against a positive diagnosis, but allow fewer than five malignant cells to be detected. Unsurprisingly, the two indeterminate designations suffer from poor inter-rater variability ^12, 17^.

There are a number of drawbacks to cytological assessments, despite improvements in screening criteria: cytology slides are far less structured than traditional histological specimens (as they are a random dispersion of cells); there is high inter-rater variability; and the workload involved often leads to cytologist exhaustion– all of these factors increase the likelihood of misclassification. Furthermore, TPS does not introduce rigorous screening criteria for urothelial cell clusters, instead mainly relying on aggregates of individual cellular estimates. Systems to automate the assessment of cytology specimens can provide more quantitative assessments of atypia, while improving reliability and reproducibility.

Advances in cytopathology vis-à-vis increased automation can bring several benefits to all stakeholders in the healthcare space ^18–22^. The adoption of computer assisted Papanicolaou (‘Pap’) test screening helped laboratories address overwhelming numbers of tests that formerly required manual screening, leading to inevitable workflow backlogs and diagnostic errors resulting from overwork. The end result of this practice was the drafting of the CLIA-88 regulations concerning cytotechnologist workload limits and the development of semi-automated Pap screening devices such as the FocalPoint™ GS and the ThinPrep ® Imaging System (TIS) ^23, 24^. The commercial success of these automated systems in the gynecologic cytology market provides a window into the possibilities of future computational applications in urine cytology ^25–33^. The factors which drove the creation of automated gynecologic cytology systems are similarly present in urine cytology: to improve clinical outcomes and integrate smoothly within the daily workflows of cytopathology laboratories. Outside of gynecologic cytology, several computational methods have been developed for cytological applications in screening cancers of varying types of specimens ^18, 34–36^. For instance, efforts have been made to screen potential malignancies in thyroid fine-needle aspirations (FNA), liquid-based lung cancer specimens, pancreaticobiliary FNA, breast lesions, and urine specimens ^37–42^.

Systems to automate cytology screening can provide more quantitative assessments of atypia while improving reliability, precision and reproducibility of findings. State-of-the-art approaches leverage deep learning, which relies on the use of artificial neural networks (ANN– inspired by the central nervous system), to construct indicators of atypia that can be formulated into diagnostic tests. For instance, Sanghvi et al. developed a semi-autonomous diagnostic decision aid for bladder cancer using a deep learning algorithm to quantify abnormal cytomorphological features ^43^. The algorithm detected urothelial cells using QuPath, urothelial clusters using density-based clustering and used convolutional neural networks for scoring cells for atypia (e.g., NC ratio, hyperchromasia, etc.). Although the effectiveness of QuPath, the scoring algorithms, and density-based clustering was not fully discussed, the study showed promising results in estimating overall atypia and could potentially improve bladder cancer screening. However, it should be noted that other studies have highlighted the limitations of QuPath in disaggregation of cells within clusters in favor of detection-based approaches, indicating a need for further refinement of the algorithm ^44–47^.

We previously developed the AutoParis system to automatically report the presence of malignant cells across cytology specimens through cross-tabulation of the degree of atypia and NC ratio for all urothelial cells in the preparation ^48^. Cross-tabulation is used to generate an Atypia Burden Score (ABS) to directly classify the specimen. The current AutoParis system operates by: 1) using connected component analysis (morphometry) and watershedding to separate individual cells from cell clusters within the specimen; 2) estimating the NC ratio of the cell using a segmentation neural network to separate the nucleus and cytoplasmic components on a pixel-by-pixel basis; 3) simultaneously assigning the cell as urothelial and recording whether the cell is atypical (atypia score) from a classifier which separates negative urothelial cells, positive urothelial cells, leukocytes, red blood cells (RBCs), debris, squamous, and crystals; and 4) generating digital images in which the cells are arranged in order of atypia, which could be helpful to pathologists. Limitations in current classification systems for urine cytology include ^20^: 1) confounding by the presence of blood, high cellularity, neobladders (abundant degenerated enterocytes) and scanning artifacts. Other previously unaccounted for cell types may also confound classifiers (e.g., polyomavirus encrusted cells conflated with positive urothelial cells, leukocytes vs. clusters of leukocytes, urothelial cells with no nucleus present, renal tubule cells) ^49^; 2) morphometry algorithms may not scale to hundreds of thousands of cells at maximal resolution; 3) density-based clustering / watershedding is likely insufficient to separate overlapping cells; 4) using a single classifier does not adequately separate the tasks of determining whether a cell is both urothelial and atypical; 6) orientation and size of cell could confound the classifier; and 7) existing graphical displays for communicating the burden of atypia are static rather than dynamic.

We set out to improve on the AutoParis classification tool by addressing the above limitations and additionally trained the models using a more expansive dataset– we dub the new tool **AutoParis-X (AP-X)**. In AutoParis-X, we addressed challenges associated with cell cluster assessment by developing an artificial intelligence tool that uses detection models to localize urothelial cells, overlapping cell boundaries, dense regions of significant overlap, and identify visual markers of urothelial atypia. By breaking clusters into their constituent architectural components, this preprocessing tool facilitates downstream association studies and predictive algorithms that incorporate quantitative cluster-level features. The cell border identification tool helped develop a more comprehensive understanding of urothelial cell cluster atypia as it pertains to bladder cancer screening. In comparison to the previous AutoParis study, which was validated on a small well-curated test set, we performed a large-scale retrospective validation of AutoParis-X on nearly 1,300 real-world specimens from internal cohorts. In this manuscript, we discuss improvements to the previous approach and its potential for real-time assessment as a mature diagnostic decision aid.

## Methods

### Specimen Collection and Slide Processing

A total of 1,303 urine specimens were collected across 140 bladder cancer patients (median of 8 specimens per patient; IQR: [8-13]) from 2008 to 2019 at Dartmouth-Hitchcock Medical Center. Forty-seven of these specimens were used to curate data for training the cell and cluster-level machine learning models (***cell and cluster-level training and validation cohort***). Four specimens were removed due to equivocal findings and/or excessive confluent cellularity. AutoParis-X was further trained and validated on 1,252 specimens after curating slide-level cell/cluster predictors (***slide-level training and validation cohorts***; see **Calculation of Cell and Cluster Slide-Level Scores**). The specimens were prepared using ThinPrep® and Papanicolaou staining before being examined microscopically ^24^. The urine slides were scanned using a Leica Aperio-AT2 scanner at 40× resolution and were stored as 70% quality SVS files representing whole slide images. The slides were manually focused (by a trained technician) on a single plane during scanning, and z-stacking was not used. Patient and slide-level characteristics from the ***slide-level training and validation cohorts*** can be found in **Table 1**. All slides were assessed by a group of five cytopathologists using TPS criteria (negative for high grade urothelial carcinoma, atypical urothelial cells, suspicious for high grade urothelial carcinoma, positive for high grade urothelial carcinoma) ^12^.

**Table 1:** Patient and Specimen Cohort Characteristics.

|  | Overall |
| --- | --- |
| Number Specimens | 1252 |
| Voided (%) | 1103 (88.1) |
| Prior History Hematuria (%) | 171 (13.7) |
| Diagnosis (%) |  |
| Negative for High Grade Urothelial Carcinoma | 810 (64.7) |
| Atypical Urothelial Cells | 296 (23.6) |
| Suspicious for High Grade Urothelial Carcinoma | 98 (7.8) |
| Positive for High Grade Urothelial Carcinoma | 48 (3.8) |
| Contains Artifact (%) | 265 (21.2) |
| Number Patients | 140 |
| Age (mean (SD)) | 71.19 (12.37) |
| Sex = M (%) | 106 (75.7) |

### Methods Overview

In this section, we summarize improvements introduced in **AutoParis-X**, which will be elaborated on in following sections. **AutoParis-X** was written using the Python programming language and neural networks were implemented using the PyTorch and Detectron2 frameworks ^50, 51^. Statistical and machine learning models were implemented in Python and R ^52–54^. A graphical overview is provided in **Figure 1**:

1. **Slide processing–** Connected components analysis to isolate individual cells and cell clusters, sped up through parallel processing ^55^.
2. **Cell border detection (BorderDet)–** Isolates cells within urothelial clusters with overlapping cytoplasmic borders through neural network detection model ^44^.
3. **Cell-Level Measures:**

a. **Morphometric measures–** Additional morphological features to improve cell-type classification and atypia estimation (e.g., size / area).
b. **Urothelial Classifier (UroNet)–** Used to filter urothelial cells from potentially conflated cell types through a convolutional neural network, which operates on images of cells and their morphometric measures ^56^– trained on an expanded dataset with more cell classes.
c. **NC ratio estimation (UroSeg)–** Estimates the NC ratio by neural network pixel-wise segmentation of background, nucleus and cytoplasm. Used as objective marker of atypia.
d. **Atypia score (AtyNet)–** For predicted urothelial cells at a particular cutoff threshold, a subjective score which incorporates multiple screening criteria (e.g., hyperchromasia, etc.) is determined using another convolutional neural network which operates on images of cells and their morphometric measures and outputs an atypia score ^48^.
4. **Cell- and Cluster-Slide-level scores–** Established through a combination of the above scoring methods, counting the number of cells/clusters in the slide with atypical morphology / cluster architecture as defined by previous works ^43, 48^. Optimal decision cutoffs for determining cellular/cluster atypia were decided using Bayesian Optimization techniques ^57^.
5. **Classifier development–** Machine learning classifier which integrates cell and cluster level scores and other demographic/specimen characteristics into an Atypia Burden Score (ABS), accounting for repeat measures by patient ^58–64^.
6. **Model interpretation–** A hierarchical logistic regression model was constructed from the machine learning model to identify important indicators of atypia, in addition to analogous univariable models. Helpful graphical displays were generated through an interactive web application ^65^.
7. **Demo–** A demo was deployed to an Amazon Web Services (AWS) server and software released through GitHub and PyPI.

**Figure 1:**
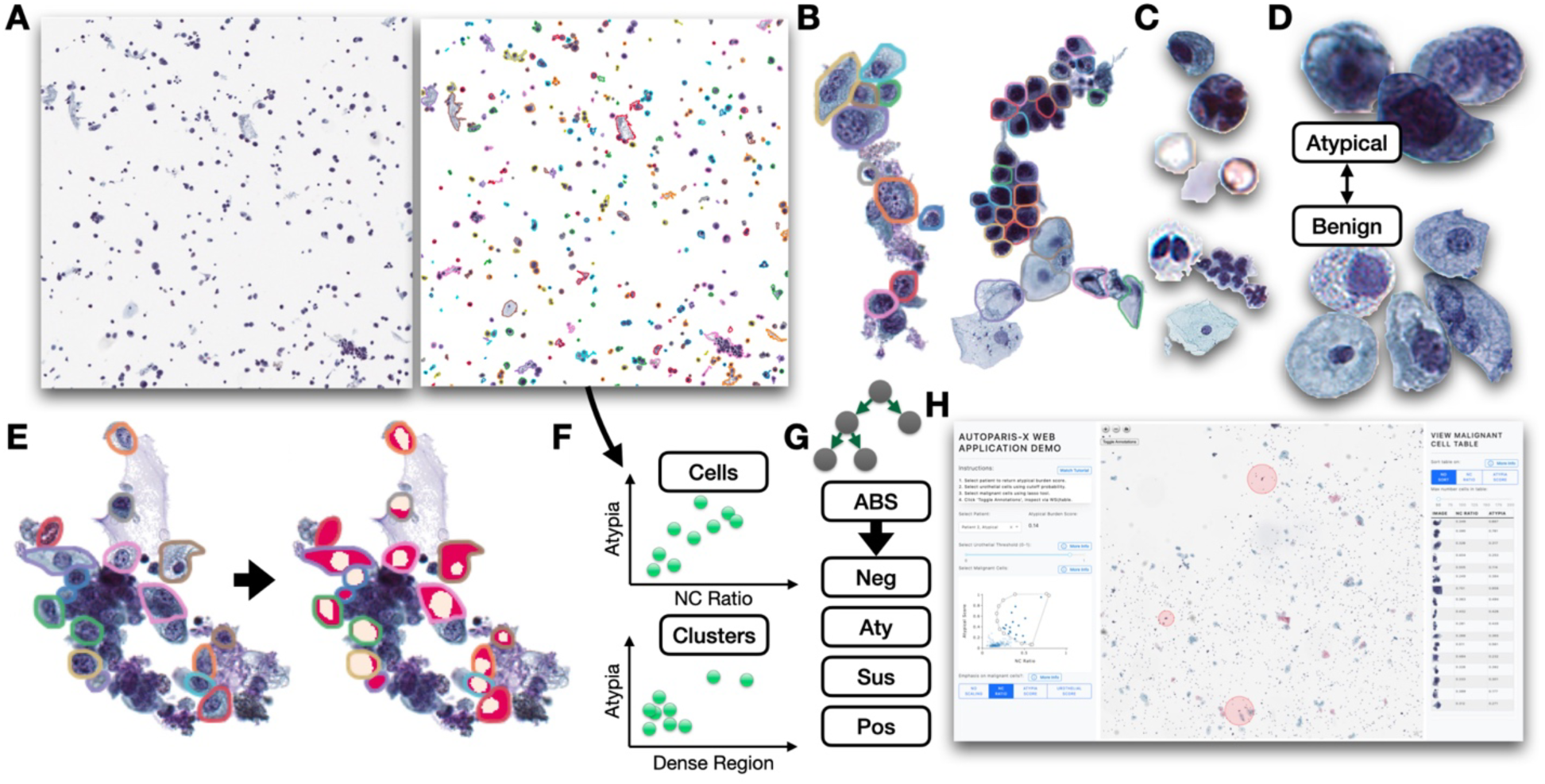
AutoParis-X specimen processing workflow: **A)** Connected component analysis isolates candidate cells and cell clusters; **B)** Individual cells and cytoplasmic borders isolated from cell clusters using BorderDet; **C)** UroNet isolates specific cell types across slide, in order: i) urothelial cells, ii) polyomavirus infected cells, iii) crystals, debris, RBCs, iv) leukocytes, v) leukocyte clusters, vi) squamous cells; **D)** AtyNet estimates atypia score for each urothelial cell; **E)** UroSeg calculates the NC ratio for each urothelial cell after being isolated using the connected component analysis or BorderDet; **F)** example rich information frame cell and cluster level scores, which cross tabulate statistics across the slide; **G)** mixed effects machine learning method predicts atypical burden score which correlates with the reported diagnosis; **H)** cytopathologists can rapidly assess the specimen using the AutoParis-X web application

### Slide Preprocessing

As detailed in a previous work, individual objects in the image were identified through a connected component analysis ^48^. In brief, WSI were converted into grey scale images using *opencv2* in Python (version 3.8) ^66^. The background of WSIs were converted to white through intensity thresholding of the grey scale image to form an object mask. Small objects, defined as a pixelwise area of 50 or below, were filtered using the *remove_small_objects* (*scipy,* Python v3.8) morphological operation ^67^. Large objects (e.g., ink markings) were similarly filtered as defined by a minimal area of 500,000 pixels. After small and large object removal, holes within the object mask were filled through the *fill_voids* function (which is faster than offerings from the *scipy* package) ^68^. We leveraged the *cupy* package (Python v3.8) to reduce compute time through usage of Graphics Processing Units (GPU) where appropriate after extensive timing tests ^69^. Subimages of slide objects (e.g., candidate urothelial cells and clusters) were returned using the *scipy regionprops* function, which also returned various other morphometric measures and bounding boxes. Inference time and memory usage for the connected component analysis for object identification was reduced through distributed computing procedures (e.g., *Dask*), which use optimized parallelization to operate on larger-than-memory arrays. Using multiprocessing through *dask*, operations were also parallelized across subregions within the slide ^55^.

### Cell Border Identification for Cell Cluster Analysis

To improve detection of individual cells within clusters, we previously developed a cell detection neural network, BorderDet, (using the state-of-the-art Detectron2 framework) to identify: 1) location of cells through estimation of bounding boxes (one box per cell) and 2) identify cell boundaries by separating overlapping cytoplasm from adjacent cells. BorderDet was developed using cell clusters identified from the ***cluster-level training cohort***. In brief, two cytopathologists (LJV and XL) annotated 800 cell cytoplasmic boundaries for squamous cells, inflammatory cells, negative/atypical urothelial cells, and dense regions of overlapping/indistinguishable cell borders (*dense region*). BorderDet is an object detection neural network that can detect multiple objects/instances (i.e., cells) in a cell cluster image ^44^. It looks for areas in the image that may contain an object and then assigns a score that indicates how likely it is that the region contains an object. The program labels identified objects with the appropriate label (e.g., squamous cell, dense region) and draws a line around the edges of the object (i.e., segmentation mask) to portray the exact boundary, which can overlap with adjacent cells. This allows the program to accurately identify and locate multiple objects in a single cluster. Objects were then filtered using non-max suppression, a technique which ranks overlapping objects, as defined through their intersection over union (IoU), based on their “objectness” score and removes objects with a lower score ^70^.

To reduce the number of objects assessed using BorderDet, a size filter was enforced, assessing candidate cell clusters with a pixelwise area of at least 1800 pixels, determined through a sensitivity analysis and visual inspection. Parallel processing through multithreading and multiprocessing was integrated using *dask* for rapid evaluation ^55^. Individual cells extracted through the connected component analysis (area between 256 and 1800 pixels) and objects extracted from clusters using their instance segmentation masks were further assessed using single-cell algorithms which report quantitative metrics of atypia (**cell-level measures**).

In comparison to the density-based clustering approach that validated urothelial clusters using a CNN (Sanghvi et al.), which could lead to many false negative findings (i.e., approach only “screens out” candidate cell clusters), urothelial cell clusters were identified by BorderDet if they contained urothelial cells ^43^. This approach improves on watershedding (AutoParis v1) and density-clustering (Sanghvi et al.) techniques as these two methods do not precisely identify cells within larger candidate clusters ^20, 43, 44, 48^. BorderDet also improves upon previous methods by locating dense urothelial cell architectures with overlapping indistinguishable cytoplasmic borders which are challenging to assess for individual cells. Furthermore, while presence of a dense architectural region in a cluster as defined by an area cutoff was used as an atypia predictor, dense architectures themselves were further subclassified as atypical if surrounding urothelial cells were labeled as atypical (as defined by morphology).

### Cellular Morphometric Measures

Various morphometric features were estimated from individual candidate cells, including: 1) area; 2) convex area; 3) eccentricity; 4) equivalent diameter; 5) extent; 6) Feret’s diameter; 7) maximum diameter; 8) filled area; 9) major axis length; 10) minor axis length; 11) perimeter; and 12) solidity, extracted using *scikit-image* (Python v3.8) ^56, 71^. These morphometric features were primarily used to help demarcate urothelial cells. As an example, urothelial cells are significantly larger than leukocytes, so cell area is an important criterion for separating the two cell types. Morphometric features were standardized using quantile transformation (implemented in *scikit-learn*, Python v3.8) within the training set to reduce the influence of any given cell on specifically which morphometric features were important for the assessment ^72^. This places greater emphasis on the imaging findings as means to delineate between different cell types.

### Urothelial Cell Classification

Urothelial cell classification was accomplished using UroNet, which was modified significantly from its original incarnation. While AutoParis estimated both the presence and atypia of the urothelial cell simultaneously ^48^, as differentiated from several other specimen constituents, AutoParis-X is chiefly focused on delineating urothelial cells from potentially conflated cell types and slide objects prior to estimating atypia. When aiming to validate the AutoParis algorithm, we noticed that a nontrivial number of urothelial cells lacked a nucleus, potentially related to being out of focus (no Z-stacking) ^73^, but were not included in our original training set and thus were often confused with other cell types with a smaller nuclear area (e.g., squamous cells). We also identified rare urothelial cells with changes consistent with a Polyomavirus cytopathic effect ^49, 74^. These cells are benign but assessment can often mimic HGUC and would certainly mislead any attempt to accurately predict the NC ratio and are thus removed by UroNet.

A total of 108,388 and 27,097 cells were manually labeled by two cytopathologists (LJV and XL) and used to train and validate the cell level model respectively from the ***cell-level training and validation cohort***. A breakdown of cell types present in this training and validation cohort is listed in **Table 2.** These cell images were combined into the following classes: 1) urothelial cells (benign/atypical), 2) urothelial cells with polyomavirus cytopathic effect, 3) debris, crystals and red blood cells (RBC), 4) leukocytes, 5) clusters of leukocytes, and 6) squamous cells. UroNet was developed using a residual neural network (ResNet18), augmented with an auxiliary layer which combines the morphometric information (e.g., area/size, eccentricity, etc.) with features extracted from ResNet18 by fusing the penultimate layer of the network with this information. The auxiliary neural network first maps the number of morphometric features, 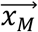, to the number of ResNet18 features using a multi-layer perceptron, *f_ϕ_*. Then the morphometric information (same dimensionality as the ResNet features) is fused with the deep learning features using a gated attention operation, which decides dynamically on a cell-by-cell basis which set of features (deep learning, 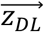, vs morphometric, 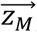 to weight more. The weight is dynamically determined using the gating neural network, *f_θ_*^75^.

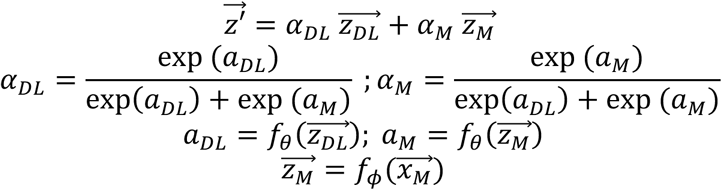

**Table 2:** Number of cell types used to train/validate UroNet and AtyNet.

|  | Benign<br>Urothelial<br>Cells | Atypical<br>Urothelial<br>Cells | Polyomavirus<br>Infected Cells | RBC | Crystals | Debris | Leukocyte | Leukocyte<br>Cluster | Squamous<br>Cells |
| --- | --- | --- | --- | --- | --- | --- | --- | --- | --- |
| **Training** | 3522 | 3795 | 3606 | 11199 | 220 | 63317 | 8037 | 3425 | 11267 |
| **Validation** | 880 | 949 | 901 | 2800 | 55 | 15830 | 2009 | 856 | 2817 |

This operation permits UroNet to filter out cells with significant size differences (e.g., leukocytes are much smaller than urothelial cells). After model training using the *PathflowAI* package ^76^, the performance of UroNet was assessed using the ***cell-level validation set*** through the area under the receiver operating characteristic curve (AUC), reported for each class. To assess how much weight was placed on the morphometric features for prediction, we investigated the attention weights, *α*, across the validation set. We used Integrated Gradients ^77, 78^, a deep learning interpretation method, to assess which specific image/deep learning and morphometric features were important for each cell type.

### NC Ratio Estimation

For cells classified as urothelial, the NC ratio was calculated for both *isolated* and *cluster cells* using a segmentation neural network, UroSeg, which employed a U-Net architecture to assign on a pixelwise basis the presence of nucleus, cytoplasm, or background ^48, 79, 80^. These areas were annotated/outlined by cytopathologists and UroSeg was trained and validated on 3,690 and 1,231 urothelial cells respectively. Performance was reported using the area under the receiver operating characteristic curve (AUC), reported on a pixelwise basis. For select cell clusters, we compared the impact of running BorderDet, followed by UroNet and UroSeg to calculate the NC ratio as compared to running UroSeg then watershedding, as was originally done by the previous AutoParis algorithm.

### Atypia Score

Several cytopathologists determined whether every urothelial cell extracted from the ***cell-level training and validation cohort*** (**Table 2**) was benign or atypical, based on existing markers of atypia (e.g., presence of nuclear membrane irregularity, abnormal chromatin, hyperchromasia, etc.). From this information, AtyNet, a CNN based on ResNet18 with a similar architecture as UroNet, was trained to recapitulate these subjective findings ^81^. For every urothelial cell, AtyNet calculates a subjective marker of atypia– the *atypia score*– which is a value from 0-1 that reflects the probability that a cell is atypical. We used IntegratedGradients, a deep learning interpretation method, to assess which specific image/deep learning and morphometric features were important for atypia assignment.

### Calculation of Cell and Cluster Slide-Level Scores

All extracted individual cell and cluster level statistics are placed into Rich Information Frames (RIF), which are data frame/tabular data structures ^48^. For any given WSI, there are three RIFs (see **Table 3** for description of features):

1. *Isolated-Cell-RIF*: Stores morphometric measures; bounding box locations within specimens, cell type assignment probabilities; NC ratios; and atypia scores for each cell not associated with clusters (*isolated urothelial cells*).
2. *Cluster-Cell-RIF:* Stores morphometric measures; bounding box locations within specimens; cell type assignment probabilities; NC ratios; and atypia scores for each cell associated with clusters, in addition to their cluster assignment label (*cluster urothelial cells*).
3. *Cluster-RIF*: Stores bounding box locations within WSI; cluster size; cytoplasmic borders; area of dense regions in cluster; and associated cluster label/identifier. Information on cellular atypia (e.g., number of atypical cells), number of urothelial cells, amongst other cluster-level measures, were added to this *RIF* from the *Cluster-Cell-RIFs*.

**Table 3:**
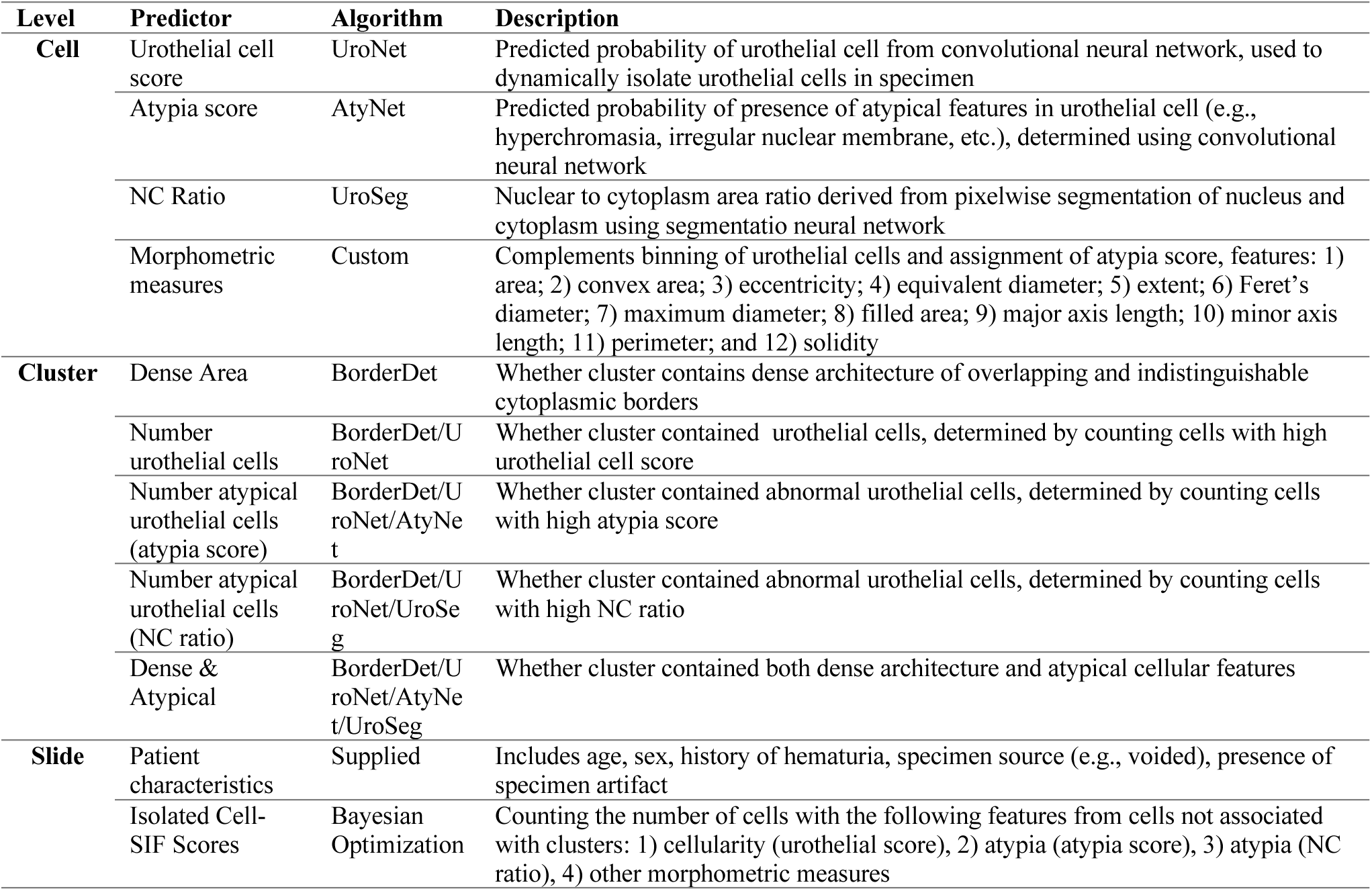
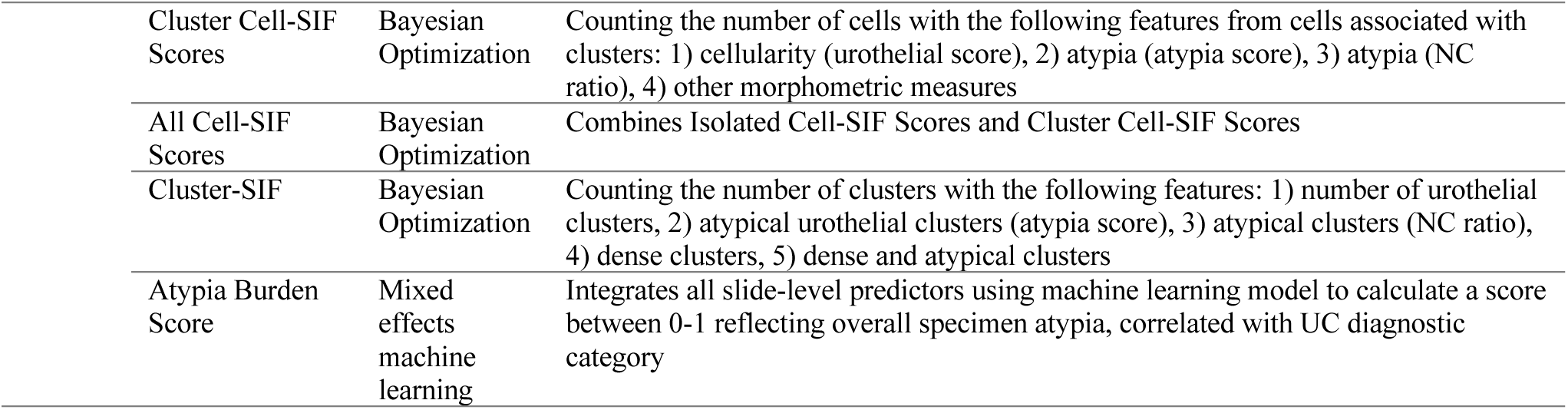
Cell/Cluster/Slide-Level Features and their descriptions.

All *RIFs* are cross-tabulated to form a Slide Inference Frame (*SIF*), which represents slide-level statistics, aggregated across all urothelial cells and urothelial cell clusters. This is accomplished by thresholding the cutoff probabilities for the cell and cluster-level scores and counting the number of cells and clusters which meet these criteria. For instance, given an atypia score cutoff of 0.7 (i.e., cell is atypical if AtyNet assigns a 70% probability), a cluster is deemed to exhibit cellular atypia if, for instance, more than 20% of the cells within the cluster are atypical under this definition. Based on the definition of a urothelial cluster (e.g., number of urothelial cells), the number of atypical clusters within the WSI can be estimated. All urothelial cells with an NC ratio of 0 were removed prior to calculating these scores. *SIF* contains the following statistics:

1. Isolated cell subscores: Derived from *Isolated-Cell-RIF*, for cells which were *not associated with clusters*, including the following statistics: 1) number of urothelial cells;
2. number of atypical urothelial cells as determined using the atypia score; 3) number of atypical urothelial cells as determined using the NC ratio; 4) number of urothelial cells; and 5) center and spread of various morphometric measures.
3. Cluster cell subscores: Derived from *Cluster-Cell-RIF*. Similar to isolated cell subscores, only considering cells which were associated with / *identified within clusters*.
4. All cell subscores: Combines isolated and cluster cell subscores, considering all cells, irrespective of whether there was a cluster assignment.
5. Cluster subscores, representing aggregate *Cluster-RIF* statistics, including: 1) number of urothelial clusters (defined by a minimum threshold of urothelial cells); 2) number of atypical urothelial clusters (defined by either NC ratio or *atypia* score); 3) number of dense clusters; and 4) number urothelial clusters that are both atypical and dense. Unlike the previous three scores which focus on individual urothelial cells, identified urothelial cell clusters represent the principal unit of analysis.

Using AutoParis-X, *RIF-SIF* scores were calculated across the ***slide-level training and validation cohorts***. We added the following patient-level characteristics to the *RIF-SIF* scores: 1) age; 2) sex; 3) history of hematuria; and 4) specimen source ^82, 83^. We also noted where slides contained significant blood, high cellularity, acellularity, neobladders (abundant degenerated enterocytes) and scanning artifacts.

### Estimating Specimen Atypia with Machine Learning

Specimen atypia was reported through dichotomization of TPS categories into the following classes: 1) negative, atypical and 2) suspicious, positive. The *Atypia Burden Score* (ABS) reflects the predicted probability of a specimen being atypical as assessed by AutoParis-X. We implemented several machine learning and statistical modeling approaches to predict specimen atypia, including: 1) generalized linear mixed effects modeling (hierarchical logistic regression; GLMM; *brms* package, R v4.1), accounting for patient- and pathologist-level random intercepts, 2) Random Forest, which does not account for clustering by patient, 3) Gaussian Process Tree Boosting (GPBoost), and 4) Bayesian Additive Regression Trees (BART) ^58–61, 64^. GPBoost and BART account for clustering by patient by fitting patient- and pathologist-level random intercepts while capturing interactions and nonlinear associations between *SIF* predictors using ensemble tree models, *f_θ_*(*x̄*):

Overall model performance was communicated using fivefold cross-validation, which randomly partitions the data into a training and validation set and reports the overall performance (using the AUC) over the validation folds. Specimens belonging to the same patient were partitioned into the same training/validation fold for each cross-validation split to avoid potential inflation of test statistics. Confidence intervals (CI) were reported using 1000-sample nonparametric bootstrapping of each fold to yield 1000 samples of cross-validation statistics. Cell and cluster-level thresholds (e.g., atypical cell if NC>0.7; atypical cluster if at least 3 urothelial cells are atypical), which are used to generate *RIF-SIF* scores, were optimally aligned with specimen atypia through a Bayesian Optimization routine ^57^.

### Interpretation

We identified significant *ABS* predictors by extracting salient interactions from the tree ensemble models and reporting odds ratios (OR) from univariable and multivariable Bayesian GLMM models: 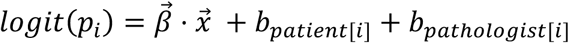. As many of the *ABS* predictors were highly multicollinear, variance inflation factors and horseshoe lasso priors were used to select predictors ^84, 85^. Univariable associations adjusting for age, sex and hematuria were reported to give credence to omitted collinear predictors in the multivariable statistical modeling.

Hierarchical Bayesian cumulative link models (i.e., ordinal regression) in a similar specification were also used to report associations between the predictors and specimen atypia, treating the urine cytology assignment as an ordinal variable ^86, 87^. Statistical significance was reported using the p-value, as derived from the probability of direction (*pd*): *p* ≈ 2 ∗ (1 − *pd*). A p-value less than 0.05 indicates a significant atypia predictor. Credible intervals, similar to confidence intervals, communicated uncertainty in the effect estimates.

### Web Application and Software Availability

We also developed an interactive web application which allows for rapid assessment of cytology slides. In brief, users first select a slide to examine. An *ABS* score is returned for the specimen as assessed using AutoParis-X. The *Cell-RIF* is converted into a 2D scatter plot of the NC ratio and atypia score– each point represents a cell. Using a “lasso tool”, users select cells within this scatterplot. The urothelial cells are highlighted on a zoomable WSI viewer (*openseadragon*) and additionally made available through an image gallery for additional examination (**Figure 2**) ^88^. The WSI viewer will highlight cells based on their relative degree of atypia as assessed algorithmically, focusing the end-user on a small subset of potentially malignant cells. A demo of this interactive web application can be found at the following URL: http://edit.autoparis.demo.levylab.host.dartmouth.edu/ (**user:** edit_user, **password:** qdp_2022; full-screen display is encouraged for optimal viewing experience). The web application also features a tutorial video for operating the application. The AutoParis-X software is also open-source, available to download on GitHub (https://github.com/jlevy44/AutoParisX) and installable using the following PyPI package: *autoparis*. Users aiming to run AutoParis-X will need to train compatible neural networks as neural networks were only trained on data from a single institution and would need additional finetuning to generalize.

**Figure 2:**
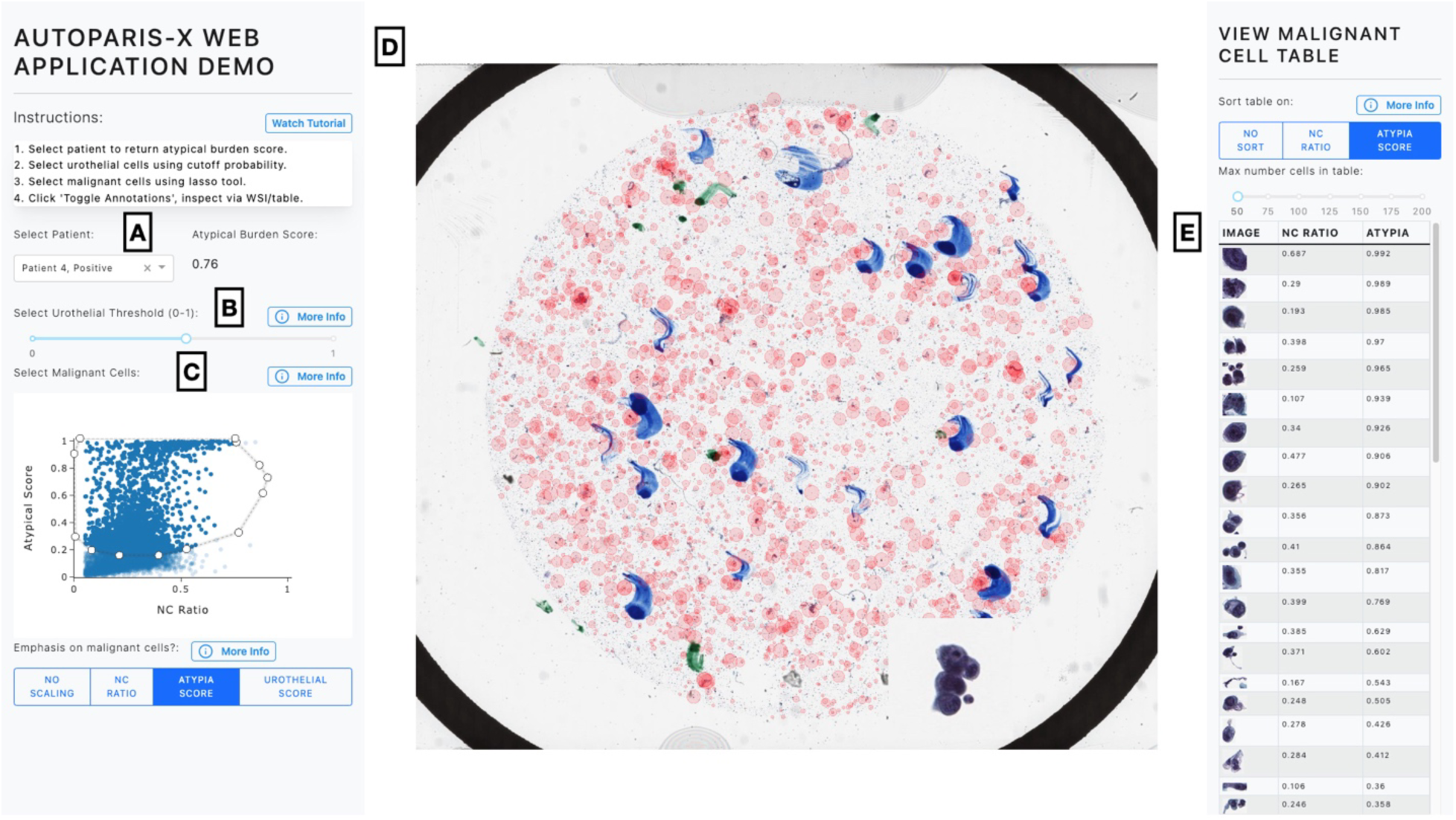
AutoParis-X Web Application: **A)** Cytopathologist selects patient/specimen scanned and processed the previous day, which outputs Atypia Burden Score; **B)** Urothelial cells are identified based on a cutoff probability selected by the user; **C)** Individual cells are plotted using scatter plot, which depicts each cell’s NC ratio and atypia score; user selects most atypical cells for viewing via the WSI viewer and gallery using the “Lasso” tool; **D)** WSI viewer– red points are sized by degree of atypia and identify important urothelial cells to assess/zoom in; **E)** gallery view enables rapid examination of individual cells, sorting them by their degree of atypia

## Results

### Performance of UroNet

UroNet demonstrated remarkable performance in the task of delineating among 6 different classes of cell types / objects to determine which cells are urothelial (**Figure 2; Table 4**). **Figure 3A** demonstrates a nearly perfect ROC curve (AUC=0.997 macro-averaged) for all 6 cell types across the validation set, indicating high classification accuracy. In addition, raw imaging features interpreted using IntegratedGradients corroborated with known histomorphology for specific cell types (e.g., highlighting dense chromatin to depict urothelial cells, surrounding membrane for squamous cells, etc.; **Figure 3B**). Many morphometric features were found to be important– for instance: 1) eccentricity as a defining feature of urothelial cells versus other cell types, 2) solidity for RBCs, 3) convex area as an important predictor for leukocyte clusters which have highly irregular formations, and 4) both convex area and solidity for squamous cells, which are larger than the other cell types and typically solid shapes without any notable deformations (**Supplementary Figure 1**). These findings suggest that UroNet can accurately identifying urothelial cells, important for establishing assessment of urothelial cells as the basis for AutoParis-X’s automated assessment.

**Figure 3:**
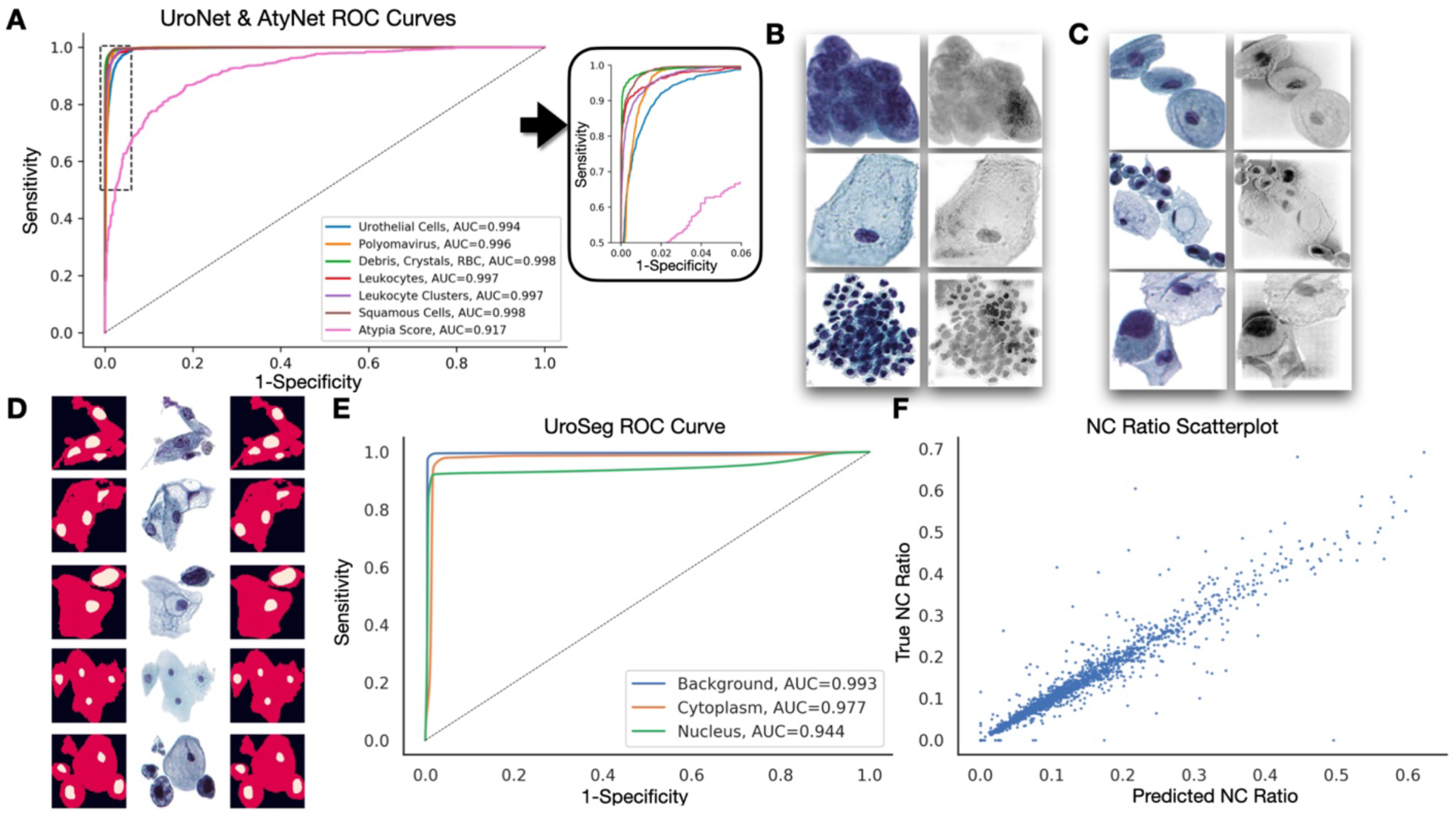
Performance of UroNet/UroSeg/AtyNet: **A)** Receiver operating characteristic curves for each cell type from the internal validation set (UroNet) and for delineating atypical versus benign urothelial cells (AtyNet); **B)** Integrated Gradients heatmap localizing important features identified using UroNet for urothelial cells, squamous cells and leukocyte clusters; **C)** Integrated Gradients heatmap localizing important features identified using AtyNet for one benign urothelial cell / cell cluster, followed by two atypical cell images; **D)** Example ground truth segmentation masks (left; background-black, cytoplasm- red, nucleus- yellow), original images (center) and segmentation masks predicted using UroSeg (right); **E)** Receiver operating characteristic curves for background, cytoplasm and nucleus (pixelwise assessments) from the internal validation set (UroSeg); **F)** Ground truth versus UroSeg predicted NC ratios, derived from the segmentation results

**Table 4:** Performance Statistics for UroNet, UroSeg, and AtyNet; 95% confidence intervals estimated using 1000-sample non-parametric bootstrapping

| Algorithm | Quantity | Measure | Estimate | 2.5% CI | 97.5% CI |
| --- | --- | --- | --- | --- | --- |
| UroNet | Urothelial | AUC | 0.994 | 0.993 | 0.995 |
|  | Polyomavirus | AUC | 0.996 | 0.995 | 0.996 |
|  | Debris, Crystals, RBCs | AUC | 0.998 | 0.998 | 0.998 |
|  | Leukocytes | AUC | 0.997 | 0.996 | 0.998 |
|  | Leukocyte Clusters | AUC | 0.997 | 0.996 | 0.997 |
|  | Squamous Cells | AUC | 0.998 | 0.998 | 0.999 |
| AtyNet | Atypia Score | AUC | 0.917 | 0.905 | 0.929 |
| UroSeg | Background | AUC | 0.993 | 0.993 | 0.993 |
|  | Cytoplasm | AUC | 0.977 | 0.977 | 0.977 |
|  | Nucleus | AUC | 0.944 | 0.944 | 0.944 |
|  | NC Ratio | Spearman | 0.965 | 0.954 | 0.973 |
|  |  | Mean Absolute Error | 0.015 | 0.014 | 0.017 |

### Performance of UroSeg

UroSeg, a neural network segmentation tool, demonstrated excellent performance on our internal validation set in predicting the pixelwise presence of the nucleus and cytoplasm (AUC=0.971 macro-averaged) in order to calculate nuclear to cytoplasm (NC) ratio (**Figures 2-3; Table 4**). **Figure 3F** also shows nearly perfect receiver operating characteristic curves for both the nucleus and cytoplasm, indicating the high accuracy of UroSeg in predicting these structures. Additionally, we found that the NC ratios calculated from the segmentation masks produced by UroSeg correlated nearly perfectly with the ground truth NC ratios (r=0.965; MAE=0.015) annotated by the cytopathologists (**Figure 3G**). **Figure 3E** demonstrates the alignment of the true and predicted nuclear and cytoplasmic segmentation masks, further highlighting the accuracy of UroSeg. UroSeg was similarly effective when used in conjunction with BorderDet, our previously established urothelial cluster border separation tool. Cells extracted from urothelial clusters using BorderDet and confirmed to be urothelial via UroNet were assessed using UroSeg. We compared the NC ratios, averaged across each urothelial cluster, in our internal validation set with what was accomplished using watershedding techniques (which divided the clusters after seeding the watershed based on the location of the nuclei). Watershedding was not sensitive to the cell type as it did not leverage BorderDet and UroNet. In addition, for clusters containing urothelial cells and background debris or other confounding cell types, watershed heavily underestimated the NC ratio (**Figure 4**). This was universal across all of the urothelial clusters in the internal validation set. Through visual examination, it is clear that by precisely demarcating cytoplasmic borders between immediately adjacent and overlapping cells, BorderDet and UroNet allow for precise estimation of the NC ratio. Opting for alternative assessment approaches (e.g., watershedding) could reduce the predictive capacity of slides containing abundance of urothelial cell clusters by removing or unnecessarily skewing the reported statistics for these cells as compared to isolated cells.

**Figure 4:**
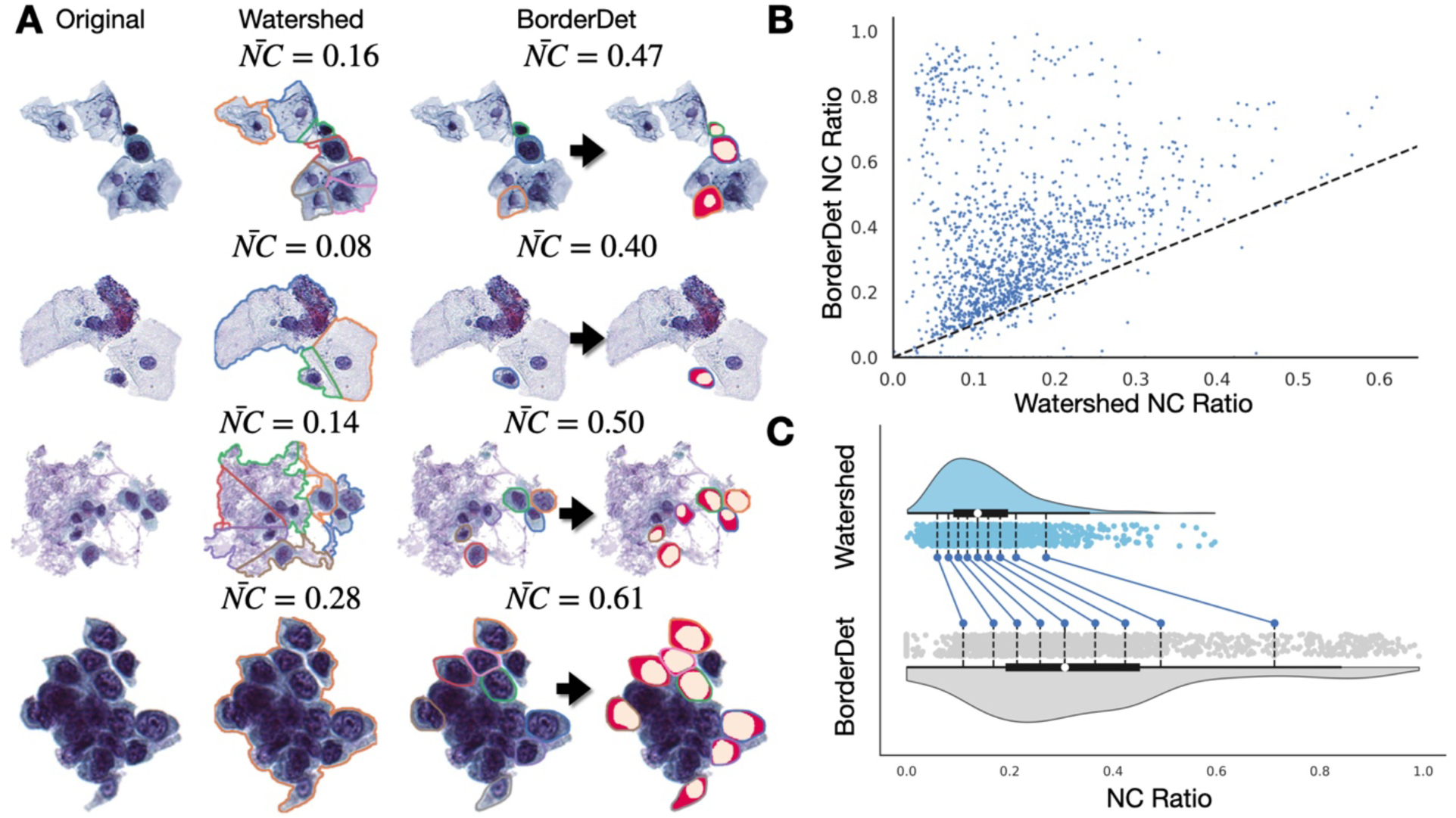
Performance of BorderDet and UroSeg on estimating NC ratios for cells in clusters: **A)** Estimates derived using watershedding underestimate the NC ratio, whereas detecting the urothelial cytoplasmic borders then using UroSeg (segmentation masks plotted over detected urothelial cells) to estimate the NC ratio leads to a higher and more accurate NC ratio; final cluster contains dense region of significantly overlapping and indistinguishable cytoplasmic borders, dense area used as a predictor for AutoParis-X; **B)** Scatterplot comparing watershed-derived and BorderDet derived NC ratios; **C)** Shift plot indicating BorderDet NC ratios are higher than that achieved using watershedding

### Performance of AtyNet

Performance for AtyNet, the neural network which provides an atypia score estimate for each urothelial cell, was equally promising (**Figure 2; Table 4**). The algorithm achieved an area under the receiver operating characteristic curve of 0.917 on the internal validation set, indicating a strong ability to distinguish between atypical and normal cells. Model interpretation using integrated gradients revealed that the algorithm placed a high emphasis on irregularities in the nuclear membrane as a key feature in determining cytological atypia (**Figure 2B**) ^56^.

### ABS Classifier Performance

Individual cell and cluster level features were cross tabulated across the slide and assessed using multiple statistical and machine learning algorithms. Many cellular and cluster level features correlated closely with specimen atypia (**Supplementary Figures 2-4**). Atypical urothelial cells as defined by both the NC ratio and atypia score, which were contained within clusters were, in some cases, more predictive of specimen atypia than assessment of isolated cells alone (e.g., cells with high NC ratio in clusters were more predictive than isolated cells with high NC ratio), further suggesting the importance of employing BorderDet for separating cells. The number of urothelial cells and cell clusters correlated directly with potential for malignancy. Urothelial cell clusters which were both atypical and contained dense regions were the third most predictive variable when assessed using univariable regression.

As part of the AutoParis-X framework, each machine learning model outputs the Atypia Burden Score (ABS)– the probability of assigning suspicious or positive UC exam as judged using AutoParis-X. Across all algorithms, ABS correlated closely with specimen atypia. The machine learning models which accounted for patient and pathologist-level variation, GPBoost and BART, outperformed all other approaches with AUCs of 0.89 and 0.88 respectively (**Figure 5A; Table 5**). The generalized linear mixed effects models also performed well. Across all models, ABS scores preserved the ordering of the UC categories (Negative<Atypical<Suspicious<Positive; **Figure 5B**). We fit an ordinal regression model to this data, which demonstrated a strong positive association with atypia (UC categories; *β* = 3.61; 95%*CI*: [3.12 − 4.11]; *p* < 0.0001). This information is corroborated by density heatmaps depicting the NC Ratio and Atypia score for individual urothelial cells across the entire cohort, after being filtered using UroNet. This yielded more than 6 million cells, which were separated based on their UC class. **Figure 5D** demonstrates the progression in cellular atypia across the categories– negative cases typically do not contain cells that have both high NC ratio and atypia, while these cells can be increasingly found at higher UC categories. Positive cases contain many cells that are both highly atypical with high NC ratio.

**Figure 5:**
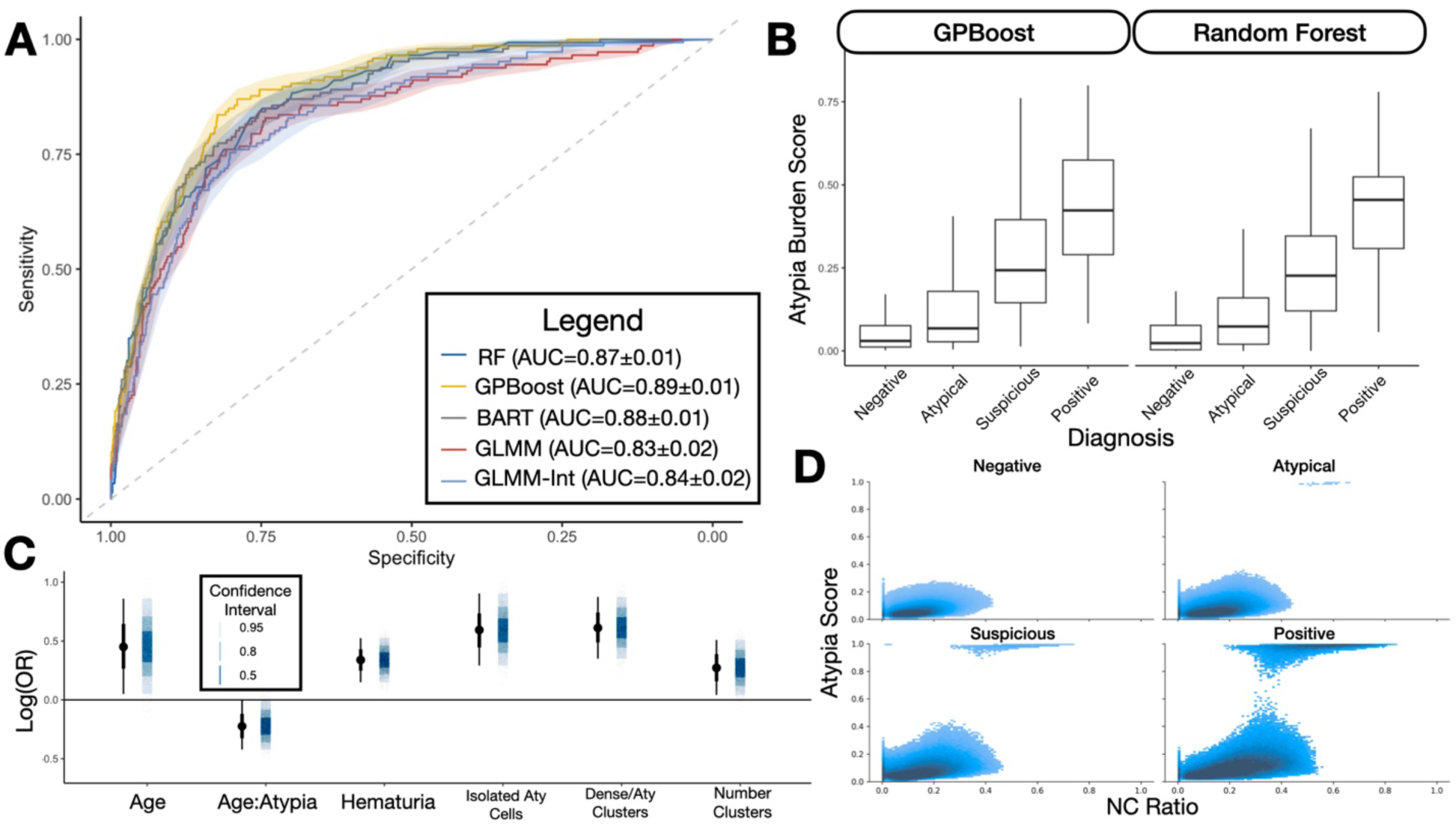
ABS Classifier Performance: **A)** Receiver operating characteristic curves illustrating performance of ABS classifiers; **B)** Boxplot of raw ABS scores predicted by GPBoost and Random Forest by UC class; **C)** Point estimates and 95% credible intervals for predictors uncovered from final multivariable Bayesian hierarchical model; **D)** Density plot of NC Ratios and Atypia scores cross tabulated across over 6 million cells from the retrospective cohort, divided by UC classes, demonstrating progression of cells to take on higher NC ratios and Atypia scores at higher UC classes

**Table 5:** Performance statistics for ABS Classifiers; 95% confidence intervals estimated using 1000-sample non-parametric bootstrapping

|  | AUC | 2.5% CI | 97.5% CI |
| --- | --- | --- | --- |
| RF | 0.873 | 0.846 | 0.897 |
| GPBoost | 0.889 | 0.866 | 0.913 |
| BART | 0.876 | 0.847 | 0.901 |
| BGLMM | 0.833 | 0.788 | 0.873 |
| BGLMM-Int | 0.843 | 0.808 | 0.874 |

### Univariable and Multivariable associations with Specimen Atypia

**Table 6** demonstrates the importance of the individual slide level predictors through both univariable and multivariable regression modeling. A few predictors remained in the unpenalized statistical model after applying the horseshoe lasso (**Figure 5C**). This included positive associations with number of clusters, number of both atypical and dense clusters, number of isolated atypical cells and an interaction between age and atypia. The interaction demonstrates that overall specimen atypia younger individuals more greatly impacted by number of atypical urothelial cells as compared to older individuals.

**Table 6:**
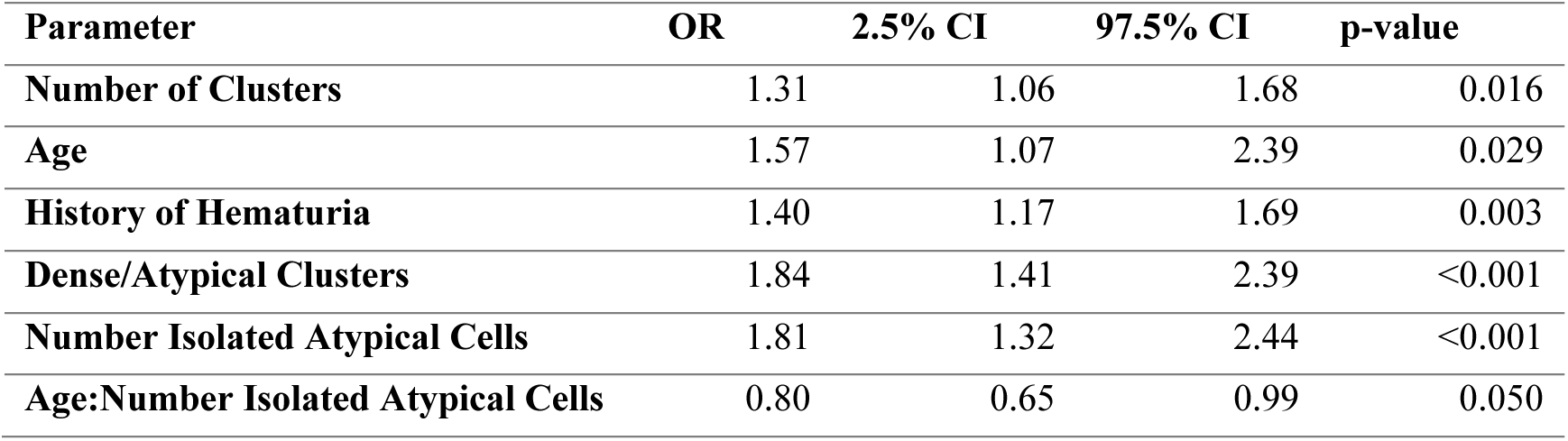
Effect estimates, 95% credible intervals and p-values for multivariable regression model.

### Web Application Example

As a demonstration of Autoparis-X’s ability to facilitate rapid examination of UC specimens, we examined four specimens with the web application (see **Supplementary Figures 5-7** for screenshots). Among thousands of specimens examined using this web tool, select cases (negative, atypical, suspicious, positive) can be further inspected using the demo application (see **Web Application and Software Availability**). The first case (**Supplementary Figure 5**) yielded an Atypia Burden Score of 0.14. Urothelial cells were selected with high atypia and were plotted on the WSI, revealing their locations. Zooming in on the WSI confirmed the reported cell-level statistics. We also used the table as means to rapidly examine all atypical cells in order of decreasing atypia as a faster method to examine cells versus zooming in using the web application. These examinations confirmed that this was in fact an atypical specimen. The second case produced an atypia burden score of 0.6– a similar examination revealed specimen atypia on par with that of a suspicious assignment. The final case was a positive patient with an atypia burden score of 0.76. We focused on only a few cells which demonstrated the highest potential for malignancy in order to focus our examination given the high cellularity of the specimen. Many of these cells were nested in urothelial cell clusters. This search identified cells which were indeed highly malignant morphologically, allowing for rapid assignment of a positive finding. In **Supplementary Figure 8**, we used the WSI viewer to zoom in on a few malignant cells identified using the AutoParis-X web application.

## Discussion

Advances in urine examination from ancient times to the information age have been accompanied by improvements in both specimen preparation and rigorous quantitative bladder cancer screening criteria ^4^. Urine cytology (UC) examination for specimen atypia has emerged as the staple of modern-day bladder cancer screening and is often accompanied by more invasive methods for cases demonstrating suspicious or positive classifications. For example, TPS is a widely used grading system in urine cytology screening for bladder cancer, which assigns four main categories based on the presence of high-grade urothelial carcinoma cells and specific cellular features. Yet, despite advances in manual examination methods, there is often poor inter-rater variability in the interpretation of atypical or suspicious specimens, and TPS does not include rigorous criteria for evaluating urothelial cell clusters ^11, 17, 89–94^. Automation in cytopathology can improve the reliability of cytological assessments and help clinicians address growing numbers of tests and avoid diagnostic errors, as has been demonstrated in the gynecologic cytology market with the adoption of systems such as ThinPrep® Imaging System and FocalPoint™ GS Imaging system ^24^. Existing systems for semi-autonomous UC examination have addressed many existing challenges, though have yet to adequately account for many additional complexities which can confound assessment (e.g., clusters, polyomavirus, etc.) ^20, 21^. In this study, we detailed the development of an artificial intelligence tool, AutoParis-X, which improves upon its previous incarnation, to allow for the rapid and nuanced examination of UC specimens; validation on a large-scale retrospective cohort illustrated the maturity and technical sophistication of this tool. For instance, challenges associated with calculation of NC ratios and overall cellular atypia within dense, overlapping urothelial cell clusters were addressed with remarkably good performance ^44^. The importance of many previously understudied predictors were evaluated (e.g., number of atypical and dense urothelial clusters). Finally, the featured interactive web application was designed for ease-of-use for semi-autonomous diagnostic decision making.

All of these innovations suggest AutoParis-X’s potential to greatly facilitate the process of bladder cancer screening, potentially resulting in a significant increase in diagnostic accuracy and a subsequent decrease in potential avenues for error (similar to what occurred with wide adoption of FocalPoint for Pap tests) ^31, 95^. For instance, results suggest that UroSeg can be used to accurately calculate NC ratios in a high-throughput manner. AutoParis-X can be used to examine hundreds to thousands of cytology specimens overnight, permitting semi-autonomous evaluation from the cytopathologist via the web application the following day (or in real time as results are generated). This is expected to increase the number and throughput of cytology exams that can be performed by any given institution while accounting for the necessary safeguards (i.e., secondary manual review of random cohort of cases as is now done with Pap tests). Cases unable to be assessed using this web-based platform could be shunted to the classical manual interpretation pathway. With any newly introduced technology, rigorous real-world clinical trials will be required to evaluate the potential impact of adopting this system. As there are only limited applications of AI technologies in digital pathology that have been approved by the FDA for clinical usage, several existing practicalities are worth addressing before AutoParis-X can be safely employed in the clinic. Social barriers for adoption can be identified through surveys on attitudes and beliefs about the tool, which will allow for iterative refinement of the output display and additional algorithmic finetuning. AutoParis-X will also need to demonstrate non-inferiority in a clinical trial (i.e., random assignment of individuals to assessment via manual and semi-autonomous examination). As non-inferiority is evaluated with respect to a ground-truth measurement, it will be difficult to prove the utility of AutoParis-X to assign specimen atypia based on alignment to cytopathologist ratings alone given the high inter-observer variation (e.g. there is no universal, quantitative ground truth in urine cytology) ^12, 17, 93^. Additional validation will likely require assessment of its capacity to predict more objective outcomes, such as disease recurrence or death ^96–99^. Additionally, its cost-effectiveness over traditional methods will also need to be proven (e.g., CPT codes, RVUs, number of specimens per day, technologist and pathologist time spent), which will communicate revenue to be expected / workforce needed when operating the device ^100–103^. A clearer understanding of how these tools can impact clinical decision making is needed before implementation (e.g., what conditions/thresholds are necessary to flag the case for manual review under a microscope) ^104^.

There are several limitations worth noting that will require future improvements and developments. We observed potential scanning artifacts (e.g., pixelation of cells), deficiencies in specimen preparation, high cellular density, and blood in the samples, which complicate the assessment. However, we have not yet developed methods to address these challenges. In addition to surveying attitudes, beliefs and adoption barriers, cytopathologists unfamiliar with digital technologies may favor assessment through analog means (e.g., microscope)– this will either require additional training and education on how to operate these nascent technologies or may require further subspecialization / training of cytopathologists to perform a digital assessment ^105–109^. AutoParis-X does not account for Z-stacking of cytology slides which can be accounted for in future iterations to model cells in 3D ^73, 110^. Annotation of individual cells and clusters were performed by a small group of cytopathologists. Some of these annotations (e.g., nucleus, delineation of cytoplasmic borders in clusters, cell type) may differ between cytopathologists. In addition, data was only collected and validated at a single institution which may limit generalization of these approaches as other institutions may have heterogenous patient characteristics/demographics and different specimen preparation methods ^111^. Additional data collection from multiple institutions can ameliorate these potential challenges by improving the diversity of the dataset, allowing additional flexibility. There is also room for improvement for deriving slide level features. While we utilized Bayesian Optimization to decide which cells/clusters were atypical, dense, clusters, etc., consideration of additional thresholds or forms to summarize this information could improve the model accuracy. There exists a plethora of modeling approaches which can be utilized to predict specimen atypia. For instance, attention and graph-based neural network architectures can take as input the entire WSI broken into constituent cells, each of which has stored attribute/morphological information. and perform what amounts to a weighted average across the cells to derive a final summary statistic ^112, 113^. The ordinal nature of UC class assignment was not explicitly taken into account for most of the results in this study and can be incorporated into these machine learning models using the appropriate model likelihoods ^114^. Institutions aiming to adopt these digital technologies will also require significant computing infrastructure. This requires the purchase and utilization of GPU enabled compute nodes (cloud computing services such as AWS and Google Cloud present viable alternatives to in-house purchases), adoption of containerized workflows, which standardize and scale analyses, and hosting of front-facing applications with appropriate databasing, security and credentialling.

## Conclusion

Bladder cancer screening through urine cytology exams is a tedious and fatigable process as cytopathologists assess tens to hundreds of thousands of cells per specimen. Algorithmic techniques to emulate these assessments are beginning to address the incredibly nuanced nature of these assessments. This study featured the design and large-scale validation of a digital diagnostic decision aid, AutoParis-X, which iterates on previous incarnations of urine cytology assessment algorithms to address many remaining complexities associated with challenging examination; further, it features a web application that allows for accurate and rapid examination of specimens. We encourage interested parties to utilize the AutoParis-X workflow and consider validating and finetuning the algorithm for other practice settings to enhance its wider generalizability. The current study demonstrated that quantitative digital urine cytology assessment methods have come of age and are prepared for further rigorous prospective evaluation to investigate its future role in augmenting clinical diagnostic decision making.

## Data Availability

Access to manuscript data is limited due to patient privacy concerns. Data produced in the present study are available upon reasonable request to the authors and four specimens are made available for demonstrated assessment at the following URL: http://edit.autoparis.demo.levylab.host.dartmouth.edu/

http://edit.autoparis.demo.levylab.host.dartmouth.edu/

## Appendix

**Supplementary Figure 1:**
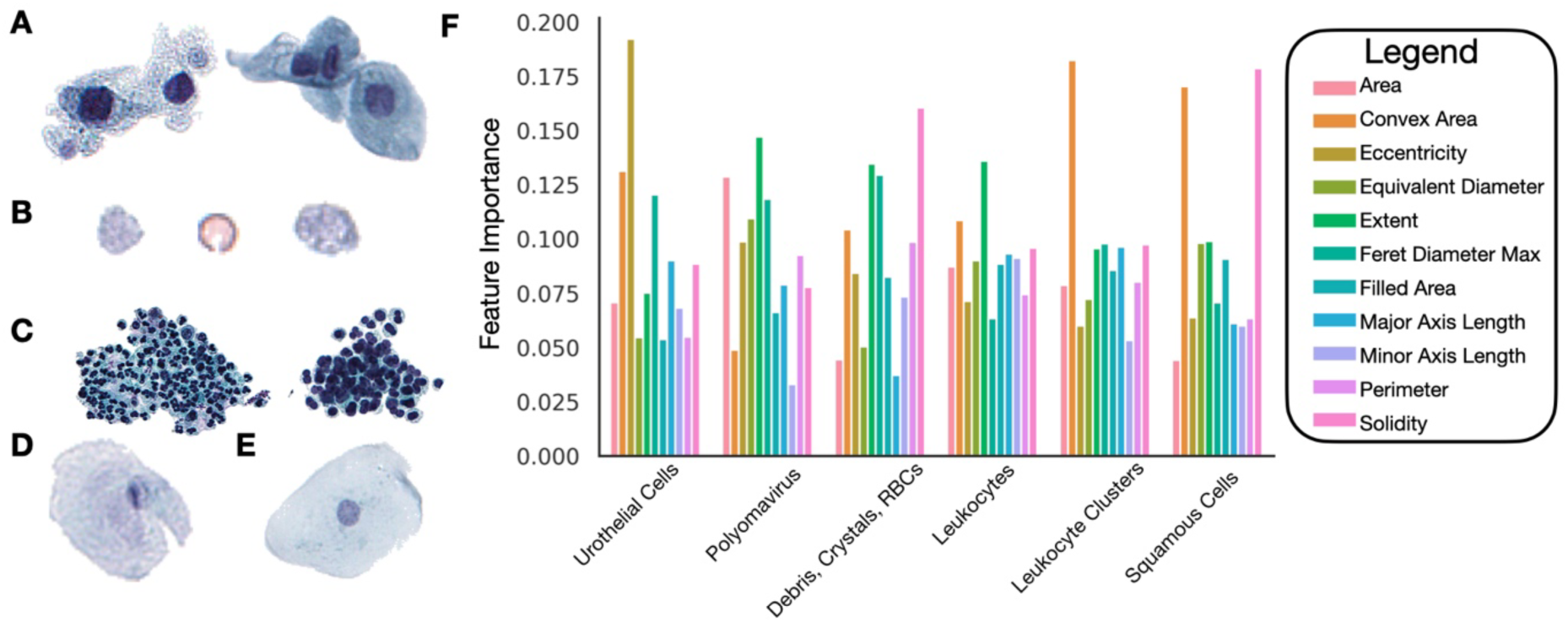
Important morphometric measures: **A)** Urothelial cells with high eccentricity; **B)** RBCs and crystals with high solidity; **C)** Leukocyte clusters with high convex area; **D)** Squamous cell with high convex area; **E)** Squamous cell with high solidity; **F)** Important morphometric features as determined using IntegratedGradients to accompany raw image features

**Supplementary Figure 2:**
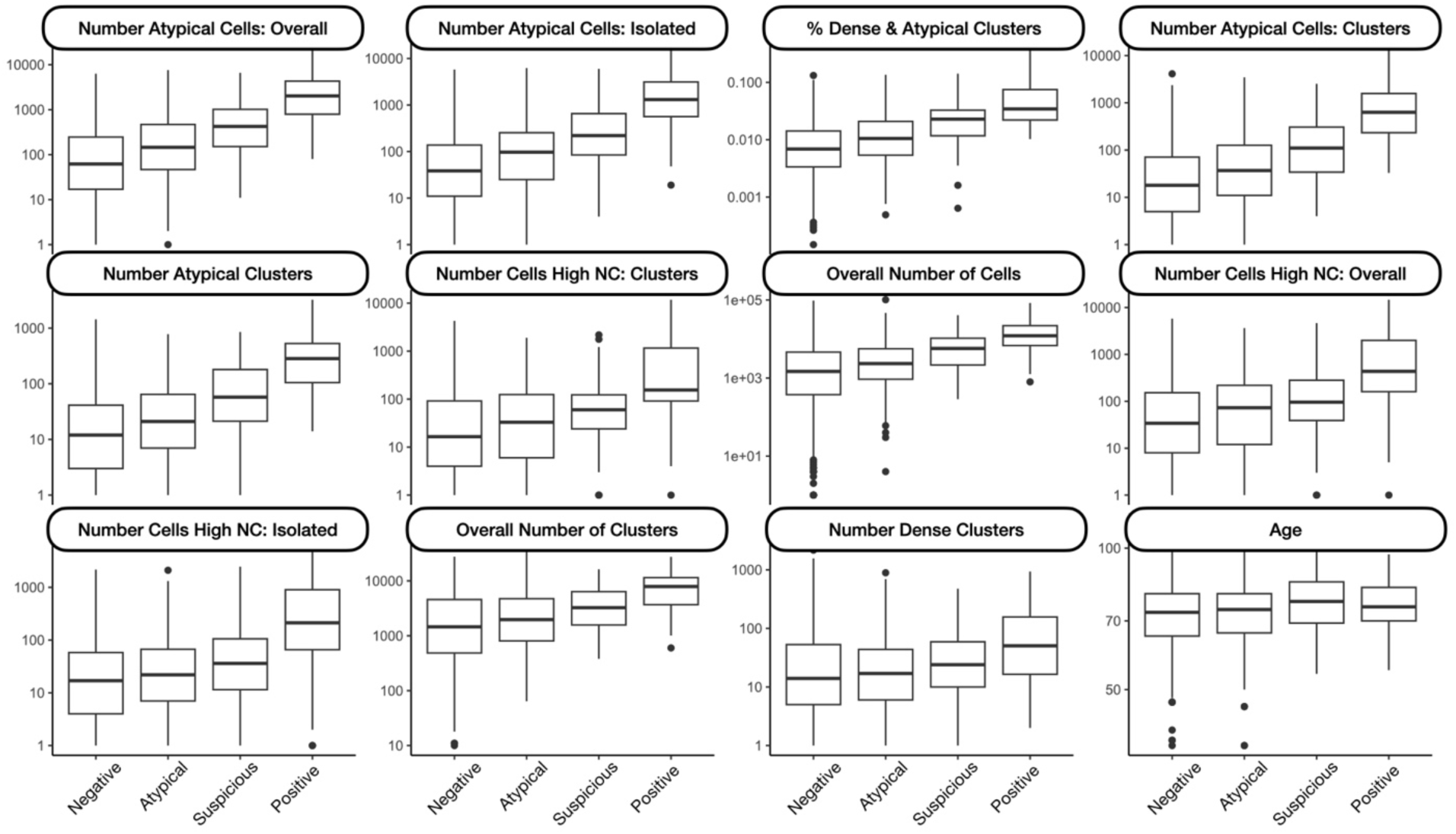
Correlation Each Slide Feature and UC Atypia, ordered by predictiveness of each feature (spearman correlation / ordinal regression)

**Supplementary Figure 3:**
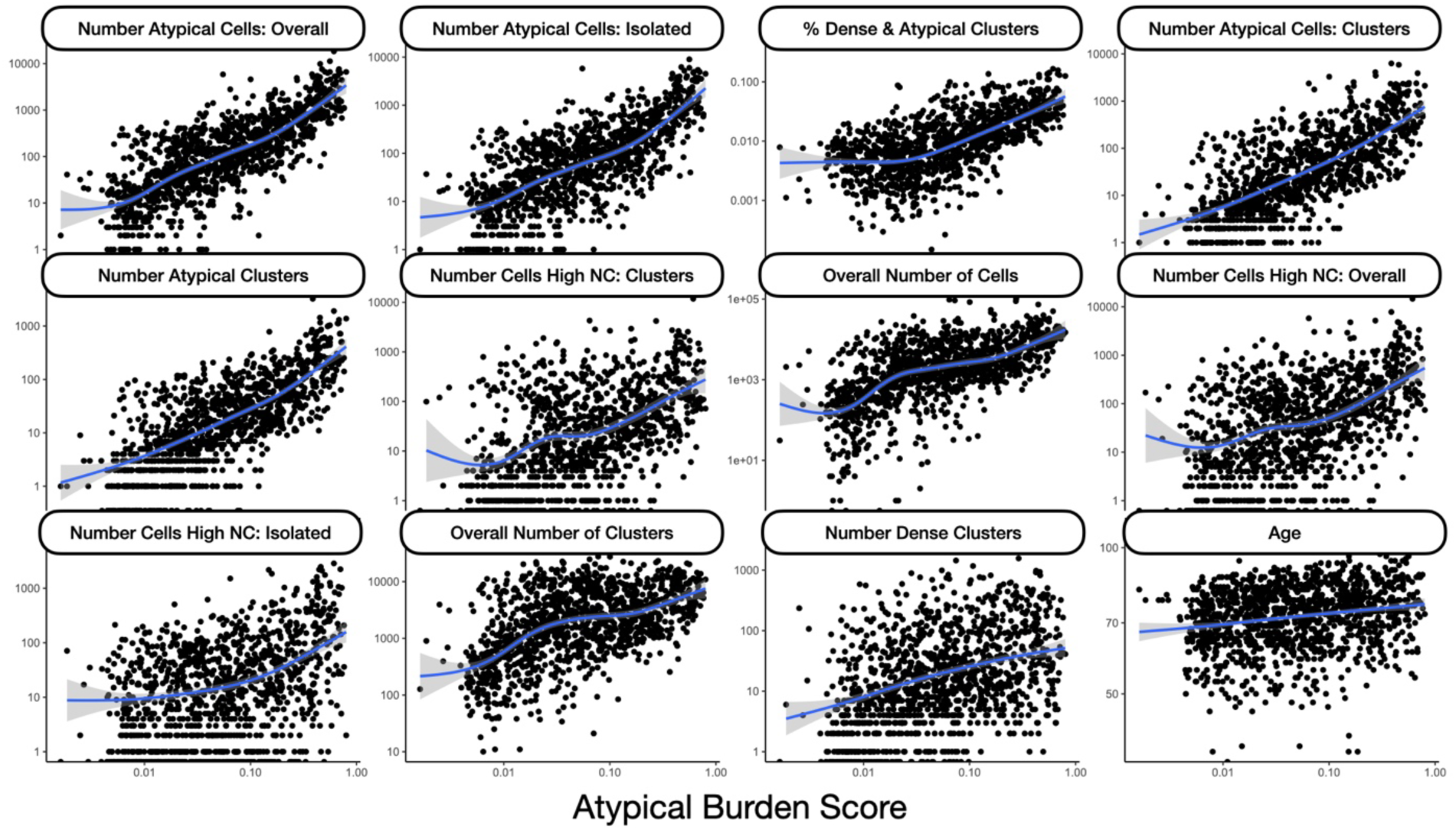
Correlation Each Slide Feature and ABS, ordered by predictiveness of each feature for UC diagnostic category (spearman correlation / ordinal regression)

**Supplementary Table 1:**
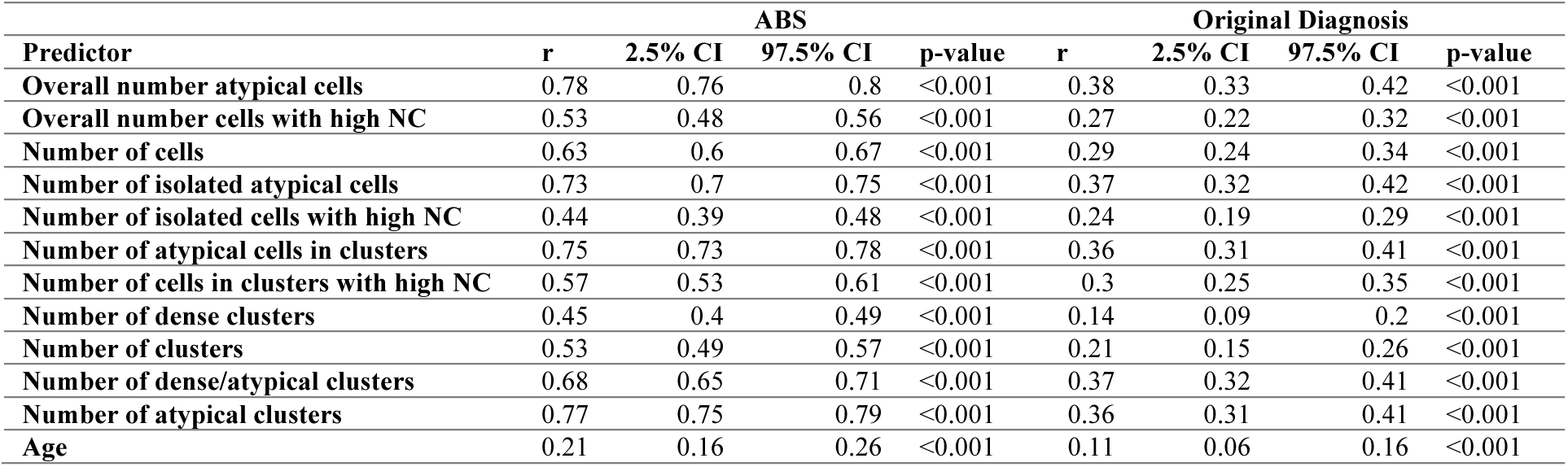
Spearman correlation between imaging predictors, ABS and original UC Class

| Predictor | ABS |  |  |  | Original Diagnosis |  |  |  |
| --- | --- | --- | --- | --- | --- | --- | --- | --- |
|  | r | 2.5% CI | 97.5% CI | p-value | r | 2.5% CI | 97.5% CI | p-value |
| Overall number atypical cells | 0.78 | 0.76 | 0.8 | <0.001 | 0.38 | 0.33 | 0.42 | <0.001 |
| Overall number cells with high NC | 0.53 | 0.48 | 0.56 | <0.001 | 0.27 | 0.22 | 0.32 | <0.001 |
| Number of cells | 0.63 | 0.6 | 0.67 | <0.001 | 0.29 | 0.24 | 0.34 | <0.001 |
| Number of isolated atypical cells | 0.73 | 0.7 | 0.75 | <0.001 | 0.37 | 0.32 | 0.42 | <0.001 |
| Number of isolated cells with high NC | 0.44 | 0.39 | 0.48 | <0.001 | 0.24 | 0.19 | 0.29 | <0.001 |
| Number of atypical cells in clusters | 0.75 | 0.73 | 0.78 | <0.001 | 0.36 | 0.31 | 0.41 | <0.001 |
| Number of cells in clusters with high NC | 0.57 | 0.53 | 0.61 | <0.001 | 0.3 | 0.25 | 0.35 | <0.001 |
| Number of dense clusters | 0.45 | 0.4 | 0.49 | <0.001 | 0.14 | 0.09 | 0.2 | <0.001 |
| Number of clusters | 0.53 | 0.49 | 0.57 | <0.001 | 0.21 | 0.15 | 0.26 | <0.001 |
| Number of dense/atypical clusters | 0.68 | 0.65 | 0.71 | <0.001 | 0.37 | 0.32 | 0.41 | <0.001 |
| Number of atypical clusters | 0.77 | 0.75 | 0.79 | <0.001 | 0.36 | 0.31 | 0.41 | <0.001 |
| Age | 0.21 | 0.16 | 0.26 | <0.001 | 0.11 | 0.06 | 0.16 | <0.001 |

**Supplementary Table 2:** Summary statistics (median, interquartile range) for each slide level feature and UC Class

|  | Overall | Negative | Atypical | Suspicious | Positive | p |
| --- | --- | --- | --- | --- | --- | --- |
| N | 1252 | 810 | 296 | 98 | 48 |  |
| Overall number atypical cells (median [IQR]) | 111.00 [24.00, 374.50] | 59.00 [16.00, 240.50] | 145.50 [46.75, 469.00] | 424.50 [151.50, 1018.50] | 2029.50 [795.50, 4322.00] | <0.001 |
| Overall number cells with high NC (median [IQR]) | 39.00 [6.00, 170.50] | 24.00 [3.00, 120.75] | 68.00 [10.00, 197.75] | 96.00 [36.00, 279.25] | 436.00 [160.50, 1995.75] | <0.001 |
| Number of cells (median [IQR]) | 2046.50 [590.25, 5856.25] | 1480.00 [370.25, 4604.25] | 2345.50 [933.50, 5629.50] | 5734.00 [2165.50, 10514.00] | 12178.00 [6810.00, 22094.75] | <0.001 |
| Number of isolated atypical cells (median [IQR]) | 57.50 [14.75, 212.25] | 36.00 [9.00, 130.25] | 94.50 [25.00, 252.75] | 220.50 [84.50, 652.00] | 1308.50 [565.25, 3127.50] | <0.001 |
| Number of isolated cells with high NC (median [IQR]) | 11.00 [1.00, 53.00] | 7.00 [1.00, 38.00] | 17.00 [3.00, 60.00] | 28.00 [6.25, 89.75] | 194.00 [51.00, 821.00] | <0.001 |
| Number of atypical cells in clusters (median [IQR]) | 24.00 [5.00, 110.25] | 15.00 [3.00, 65.00] | 36.00 [10.00, 123.75] | 110.50 [34.25, 306.75] | 631.50 [236.75, 1578.50] | <0.001 |
| Number of cells in clusters with high NC (median [IQR]) | 16.00 [2.00, 94.25] | 9.00 [1.00, 53.00] | 29.00 [5.00, 118.25] | 59.00 [23.25, 122.00] | 155.00 [92.25, 1159.25] | <0.001 |
| Number of dense clusters (median [IQR]) | 14.00 [4.00, 50.00] | 11.00 [3.00, 45.75] | 15.00 [4.75, 40.25] | 22.50 [9.25, 59.00] | 51.00 [16.50, 157.00] | <0.001 |
| Number of clusters (median [IQR]) | 1838.50 [674.75, 5228.50] | 1455.50 [486.25, 4579.75] | 1974.00 [807.50, 4744.25] | 3255.00 [1574.25, 6367.25] | 7917.50 [3695.50, 11501.00] | <0.001 |
| % clusters dense/atypical (median [IQR]) | 0.01 [0.00, 0.02] | 0.01 [0.00, 0.01] | 0.01 [0.01, 0.02] | 0.02 [0.01, 0.03] | 0.03 [0.02, 0.07] | <0.001 |
| Number of atypical clusters (median [IQR]) | 14.50 [3.00, 54.25] | 9.00 [2.00, 36.00] | 20.00 [6.00, 62.25] | 57.50 [21.25, 180.75] | 283.50 [105.50, 530.50] | <0.001 |
| ABS (median [IQR]) | 0.05 [0.02, 0.15] | 0.03 [0.01, 0.08] | 0.07 [0.03, 0.18] | 0.24 [0.15, 0.40] | 0.42 [0.29, 0.57] | <0.001 |

**Supplementary Figure 4:**
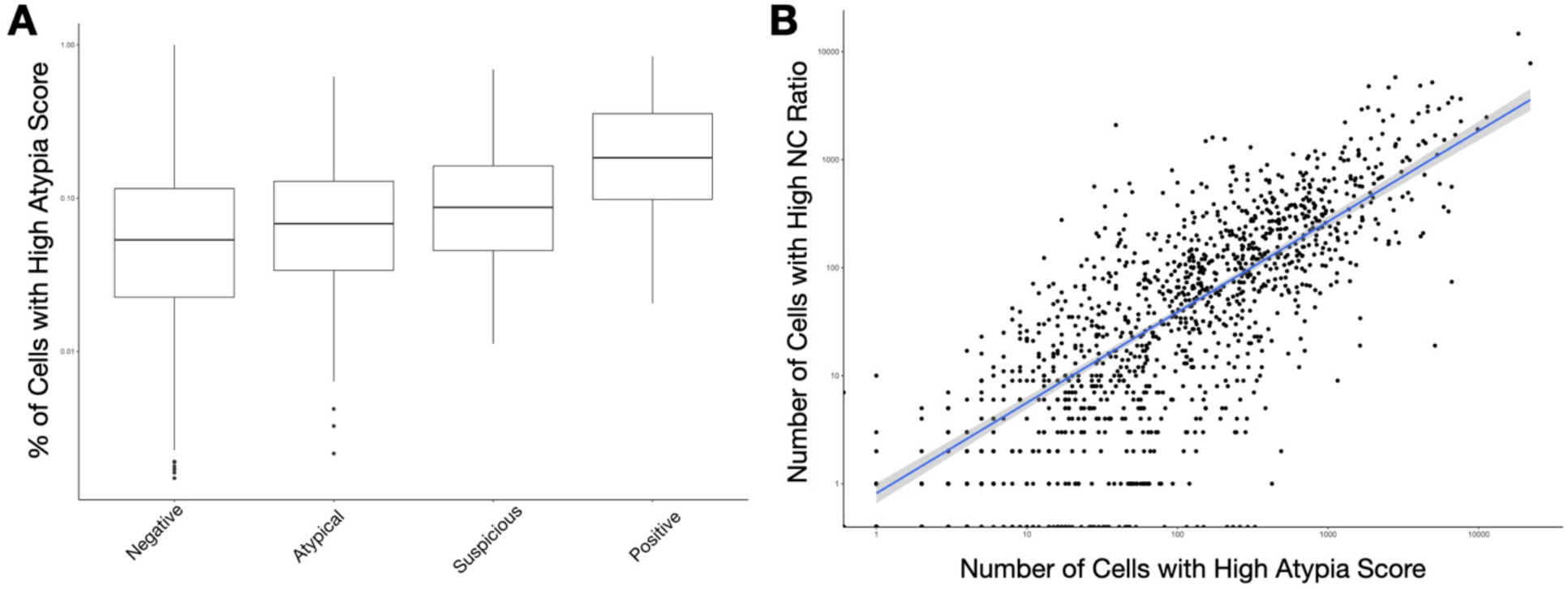
Additional associations with specimen atypia: **A)** Boxplots depicting correlation between the percentage of urothelial cells with high atypia and UC Class**; B)** Scatterplot demonstrating correlation between slide level atypia via number of cells with high atypia and high NC ratio

**Supplementary Figure 5:**
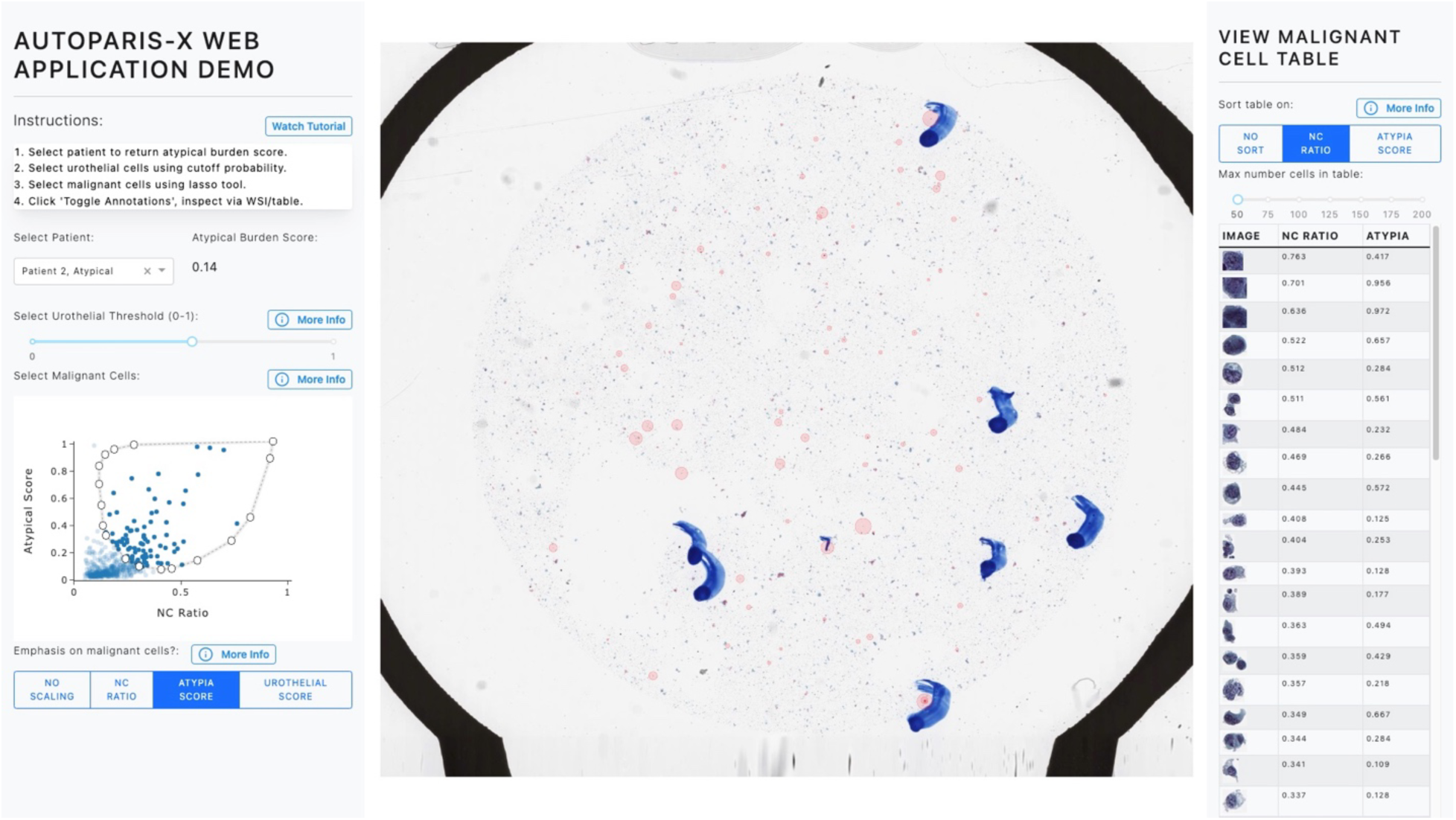
Example of identifying malignant cells in atypical slide.

**Supplementary Figure 6:**
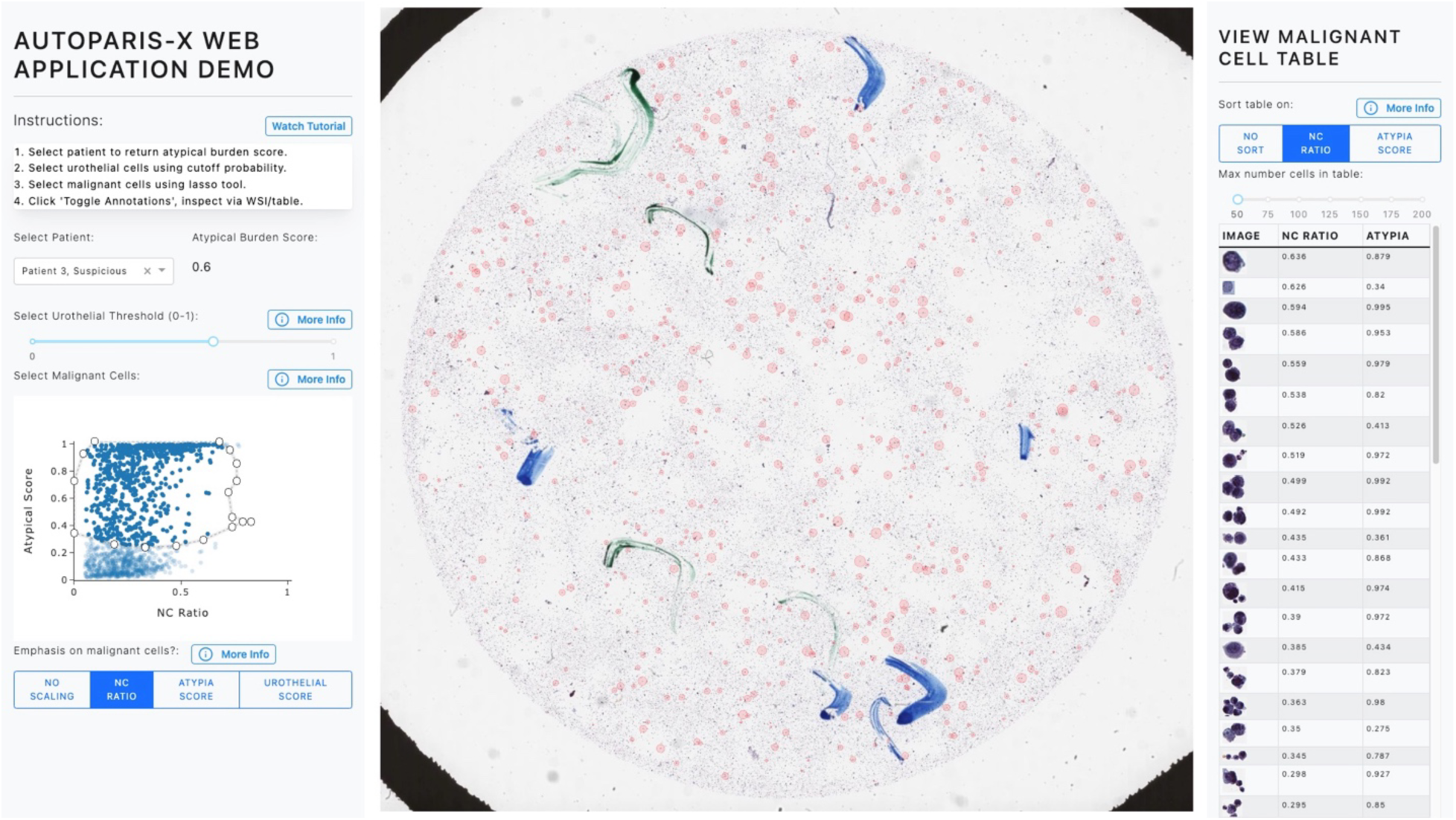
Example of identifying malignant cells in suspicious slide.

**Supplementary Figure 7:**
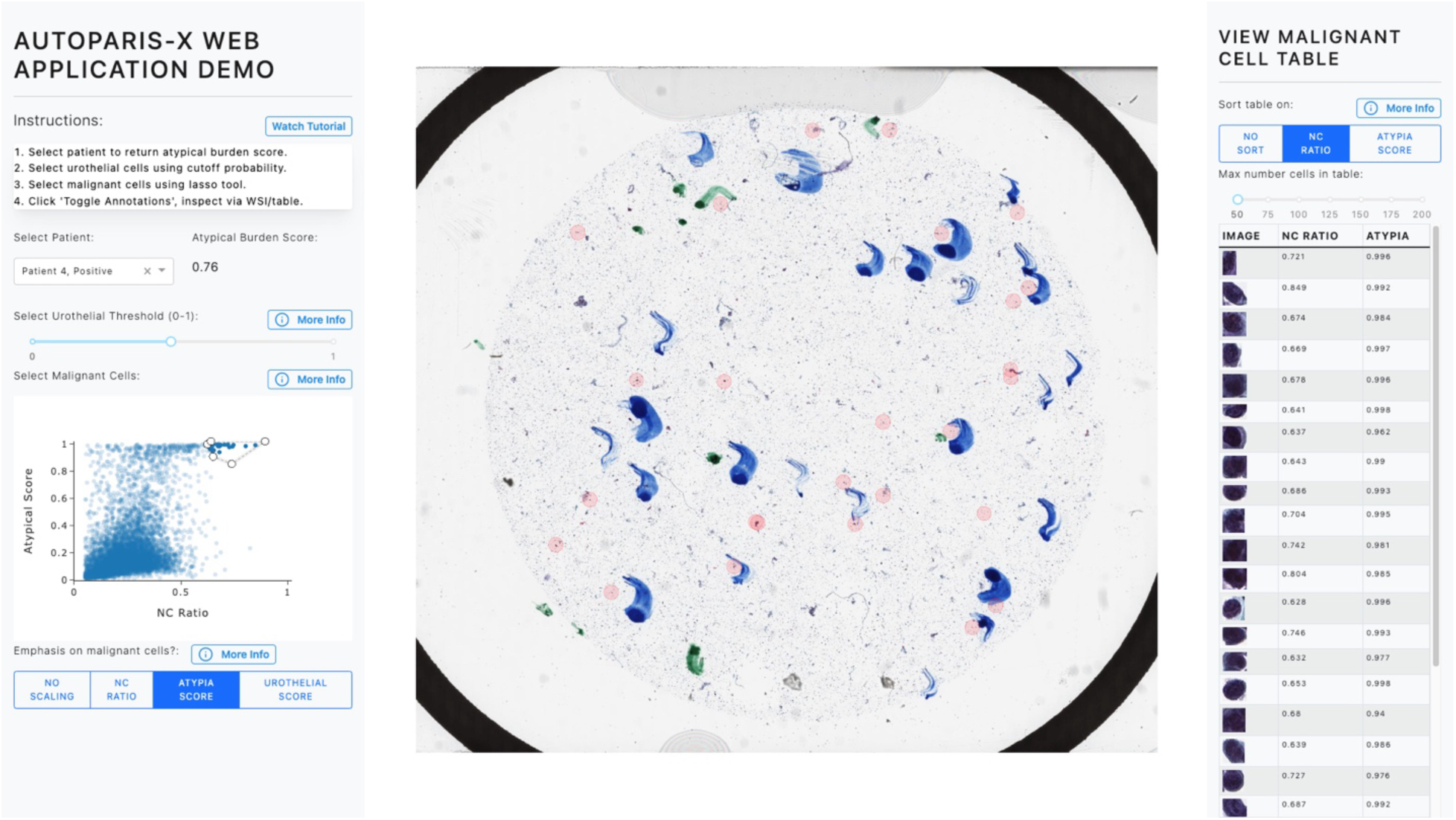
Example of identifying malignant cells in positive slide, only focusing on those with high atypia

**Supplementary Figure 8:**
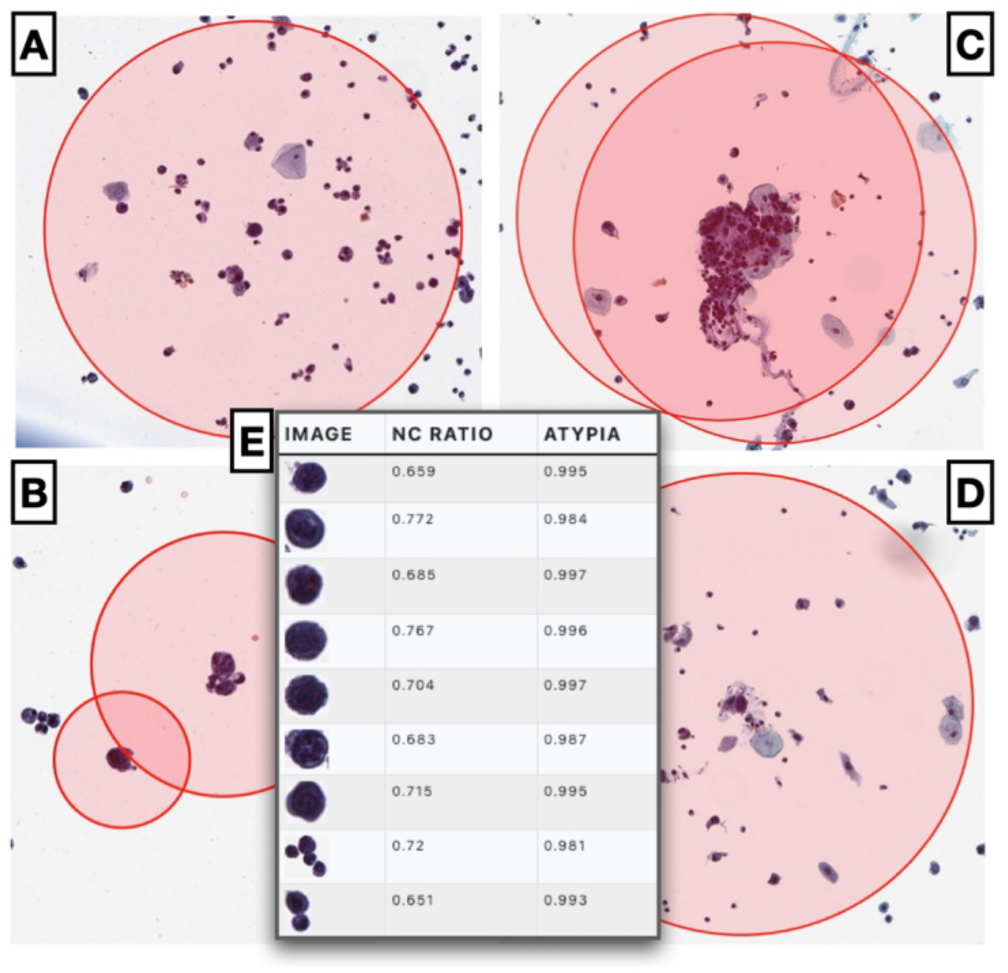
Example of atypical cells identified using Autoparis-X web application within demonstration on example atypical/positive slides: A) Isolated cell, B) Two cells with differing atypia; cell with larger red dot has higher atypia, C-D) Focusing on specific cells identified using BorderDet in hard-to-separate clusters; E) Example table of malignant cells with reported atypia scores from suspicious case

## References

1. Barkan, G. A. et al. The Paris System for Reporting Urinary Cytology: The Quest to Develop a Standardized Terminology. ACY 60, 185–197 (2016).

2. Bostwick, D. G. 7 - Urine Cytology. in Urologic Surgical Pathology (Fourth Edition) (eds. Cheng, L., MacLennan, G. T. & Bostwick, D. G.) 322–357.e7 (Elsevier, 2020).

3. Mossanen, M. & Gore, J. L. The burden of bladder cancer care: direct and indirect costs. Curr Opin Urol 24, 487–491 (2014).

4. Magiorkinis, E. & Diamantis, A. The fascinating story of urine examination: From uroscopy to the era of microscopy and beyond. Diagnostic Cytopathology 43, 1020–1036 (2015).

5. Salem, S., Mitchell, R. E., El-Alim El-Dorey, A., Smith, J. A. & Barocas, D. A. Successful control of schistosomiasis and the changing epidemiology of bladder cancer in Egypt. BJU international 107, 206–211 (2011).

6. Botelho, M. C., Alves, H. & Richter, J. Halting Schistosoma haematobium-associated bladder cancer. International journal of cancer management 10, (2017).

7. Papanicolaou, G. N. Cytology of the urine sediment in neoplasms of the urinary tract. The Journal of urology 57, 375–379 (1947).

8. Layfield, L. J., Elsheikh, T. M., Fili, A., Nayar, R. & Shidham, V. Review of the state of the art and recommendations of the Papanicolaou Society of Cytopathology for urinary cytology procedures and reporting: the Papanicolaou Society of Cytopathology Practice Guidelines Task Force. Diagnostic cytopathology 30, 24–30 (2004).

9. Wang, Y.-H. et al. Diagnostic Agreement for High-Grade Urothelial Cell Carcinoma in Atypical Urine Cytology: A Nationwide Survey Reveals a Tendency for Overestimation in Specimens with an N/C Ratio Approaching 0.5. Cancers 12, 272 (2020).

10. Barkan, G. A. Enough is enough: adequacy of voided urine cytology. (2016).

11. Roy, M. et al. An institutional experience with The Paris System: A paradigm shift from ambiguous terminology to more objective criteria for reporting urine cytology. Cytopathology 28, 509–515 (2017).

12. Levy, J. J. et al. Large-scale longitudinal comparison of urine cytological classification systems reveals potential early adoption of The Paris System criteria. J Am Soc Cytopathol S2213-2945(22)00241–1 (2022) doi:10.1016/j.jasc.2022.08.001.

13. Kurtycz, D. F., Wojcik, E. M. & Rosenthal, D. L. Perceptions of Paris: an international survey in preparation for The Paris System for Reporting Urinary Cytology 2.0 (TPS 2.0). Journal of the American Society of Cytopathology 12, 66–74 (2023).

14. Nikas, I. P. et al. The Paris System for Reporting Urinary Cytology: A Meta-Analysis. Journal of Personalized Medicine 12, 170 (2022).

15. Wojcik, E. M., Kurtycz, D. F. & Rosenthal, D. L. The Paris system for reporting urinary cytology. (Springer, 2022).

16. Wojcik, E. M., Kurtycz, D. F. & Rosenthal, D. L. We’ll always have Paris the Paris system for reporting urinary cytology 2022. Journal of the American Society of Cytopathology 11, 62–66 (2022).

17. Long, T. et al. Interobserver reproducibility of The Paris System for Reporting Urinary Cytology. Cytojournal 14, 17 (2017).

18. Landau, M. S. & Pantanowitz, L. Artificial intelligence in cytopathology: a review of the literature and overview of commercial landscape. Journal of the American Society of Cytopathology 8, 230–241 (2019).

19. Pouliakis, A. et al. Artificial Neural Networks as Decision Support Tools in Cytopathology: Past, Present, and Future. Biomed Eng Comput Biol 7, 1–18 (2016).

20. McAlpine, E. D., Pantanowitz, L. & Michelow, P. M. Challenges Developing Deep Learning Algorithms in Cytology. ACY 65, 301–309 (2021).

21. Thiryayi, S. A. & Rana, D. N. Urine cytopathology: challenges, pitfalls, and mimics. Diagnostic Cytopathology 40, 1019–1034 (2012).

22. Jiang, H. et al. Deep learning for computational cytology: A survey. Med Image Anal 84, 102691 (2023).

23. Rezende, M. T., Bianchi, A. G. C. & Carneiro, C. M. Cervical cancer: Automation of Pap test screening. Diagn Cytopathol 49, 559–574 (2021).

24. Okuda, C. et al. Quantitative cytomorphological comparison of SurePath and ThinPrep liquid-based cytology using high-grade urothelial carcinoma cells. Cytopathology 32, 654– 659 (2021).

25. Tolles, W. E. The cytoanalyzer-an example of physics in medical research. Trans N Y Acad Sci 17, 250–256 (1955).

26. Bourghardt, S., Hyden, H. & Nyquist, B. A scanning and computing microphotometer for cell analyses. Experientia 11, 163–165 (1955).

27. Abels, E. et al. Computational pathology definitions, best practices, and recommendations for regulatory guidance: a white paper from the Digital Pathology Association. J Pathol 249, 286–294 (2019).

28. Pantanowitz, L. Improving the Pap test with artificial intelligence. Cancer Cytopathol 130, 402–404 (2022).

29. Xue, P. et al. Deep learning in image-based breast and cervical cancer detection: a systematic review and meta-analysis. NPJ Digit Med 5, 19 (2022).

30. Hou, X. et al. Artificial Intelligence in Cervical Cancer Screening and Diagnosis. Front Oncol 12, 851367 (2022).

31. Thrall, M. J. Automated screening of Papanicolaou tests: A review of the literature. Diagn Cytopathol 47, 20–27 (2019).

32. Chantziantoniou, N. BestCyte® Cell Sorter Imaging System: Primary and adjudicative whole slide image rescreening review times of 500 ThinPrep Pap test thin-layers - An intra-observer, time-surrogate analysis of diagnostic confidence potentialities. J Pathol Inform 13, 100095 (2022).

33. Delga, A. et al. Evaluation of CellSolutions BestPrep® automated thin-layer liquid-based cytology Papanicolaou slide preparation and BestCyte® cell sorter imaging system. Acta Cytol 58, 469–477 (2014).

34. Pantanowitz, L. & Bui, M. M. Image Analysis in Cytopathology. in Monographs in Clinical Cytology (eds. Bui, M. M. & Pantanowitz, L.) vol. 25 91–98 (S. Karger AG, 2020).

35. Pantanowitz, L., Hornish, M. & Goulart, R. A. Informatics applied to cytology. Cytojournal 5, 16 (2008).

36. Wilbur, D. C. Digital cytology: current state of the art and prospects for the future. Acta Cytol 55, 227–238 (2011).

37. Dov, D. et al. Weakly supervised instance learning for thyroid malignancy prediction from whole slide cytopathology images. Med Image Anal 67, 101814 (2021).

38. Yao, K. et al. A Study of Thyroid Fine Needle Aspiration of Follicular Adenoma in the ‘Atypia of Undetermined Significance’ Bethesda Category Using Digital Image Analysis. J Pathol Inform 13, 100004 (2022).

39. Girolami, I. et al. Impact of image analysis and artificial intelligence in thyroid pathology, with particular reference to cytological aspects. Cytopathology 31, 432–444 (2020).

40. Sanyal, P., Mukherjee, T., Barui, S., Das, A. & Gangopadhyay, P. Artificial Intelligence in Cytopathology: A Neural Network to Identify Papillary Carcinoma on Thyroid Fine-Needle Aspiration Cytology Smears. J Pathol Inform 9, 43 (2018).

41. Elliott Range, D. D., et al. Application of a machine learning algorithm to predict malignancy in thyroid cytopathology. Cancer Cytopathol 128, 287–295 (2020).

42. Guan, Q. et al. Deep convolutional neural network VGG-16 model for differential diagnosing of papillary thyroid carcinomas in cytological images: a pilot study. J Cancer 10, 4876–4882 (2019).

43. Sanghvi, A. B., Allen, E. Z., Callenberg, K. M. & Pantanowitz, L. Performance of an artificial intelligence algorithm for reporting urine cytopathology. Cancer Cytopathology 127, 658–666 (2019).

44. Levy, J. J. et al. Uncovering additional predictors of urothelial carcinoma from voided urothelial cell clusters through a deep learning-based image preprocessing technique. Cancer Cytopathol (2022) doi:10.1002/cncy.22633.

45. Mahmood, F. et al. Deep Adversarial Training for Multi-Organ Nuclei Segmentation in Histopathology Images. IEEE Transactions on Medical Imaging (2020) doi:10.1109/TMI.2019.2927182.

46. Bankhead, P. et al. QuPath: Open source software for digital pathology image analysis. Sci Rep 7, 1–7 (2017).

47. Humphries, M. P., Maxwell, P. & Salto-Tellez, M. QuPath: The global impact of an open source digital pathology system. Computational and Structural Biotechnology Journal 19, 852–859 (2021).

48. Vaickus, L. J., Suriawinata, A. A., Wei, J. W. & Liu, X. Automating the Paris System for urine cytopathology—A hybrid deep-learning and morphometric approach. Cancer Cytopathology 127, 98–115 (2019).

49. Singh, H. K., Bubendorf, L., Mihatsch, M. J., Drachenberg, C. B. & Nickeleit, V. Urine Cytology Findings of Polyomavirus Infections. in Polyomaviruses and Human Diseases (ed. Ahsan, N.) 201–212 (Springer, 2006). doi:10.1007/0-387-32957-9_15.

50. Paszke, A. et al. PyTorch: An Imperative Style, High-Performance Deep Learning Library. arXiv:1912.01703 [cs, stat] (2019).

51. Wu, Y., Kirillov, A., Massa, F., Lo, W.-Y. & Girshick, R. Detectron2. (2019).

52. Matthes, E. Python Crash Course, 2nd Edition: A Hands-On, Project-Based Introduction to Programming. (No Starch Press, 2019).

53. Bürkner, P.-C. brms: An R Package for Bayesian Multilevel Models Using Stan. Journal of Statistical Software 80, 1–28 (2017).

54. Tippmann, S. Programming tools: Adventures with R. Nature 517, 109–110 (2015).

55. Rocklin, M. Dask: Parallel Computation with Blocked algorithms and Task Scheduling. in 126–132 (2015). doi:10.25080/Majora-7b98e3ed-013.

56. Harvey, S. E. & VandenBussche, C. J. Nuclear membrane irregularity in high-grade urothelial carcinoma cells can be measured by using circularity and solidity as morphometric shape definitions in digital image analysis of urinary tract cytology specimens. Cancer Cytopathol (2023) doi:10.1002/cncy.22682.

57. Louppe, G. Bayesian optimisation with scikit-optimize. in PyData Amsterdam (2017).

58. Sigrist, F. Gaussian Process Boosting. Journal of Machine Learning Research 23, 1–46 (2022).

59. Sigrist, F. Latent Gaussian Model Boosting. IEEE Transactions on Pattern Analysis and Machine Intelligence 1–1 (2022) doi:10.1109/TPAMI.2022.3168152.

60. Tan, Y. V. & Roy, J. Bayesian additive regression trees and the General BART model. Statistics in Medicine 38, 5048–5069 (2019).

61. Chipman, H. A., George, E. I. & McCulloch, R. E. BART: Bayesian additive regression trees. The Annals of Applied Statistics 4, 266–298 (2010).

62. Hajjem, A., Bellavance, F. & Larocque, D. Mixed-effects random forest for clustered data. Journal of Statistical Computation and Simulation 84, 1313–1328 (2014).

63. Levy, J. J. et al. Mixed Effects Machine Learning Models for Colon Cancer Metastasis Prediction using Spatially Localized Immuno-Oncology Markers. Pac Symp Biocomput 27, 175–186 (2022).

64. Bürkner, P.-C. Advanced Bayesian Multilevel Modeling with the R Package brms. The R Journal 10, 395–411 (2018).

65. Perkel, J. M. Data visualization tools drive interactivity and reproducibility in online publishing. Nature 554, 133–134 (2018).

66. Bradski, G. The openCV library. Dr. Dobb’s Journal: Software Tools for the Professional Programmer 25, 120–123 (2000).

67. Virtanen, P. et al. SciPy 1.0: fundamental algorithms for scientific computing in Python. Nature methods 17, 261–272 (2020).

68. Silversmith, W. fill-voids: Fill voids in 3D binary images fast.

69. Nishino, R. & Loomis, S. H. C. Cupy: A numpy-compatible library for nvidia gpu calculations. 31st confernce on neural information processing systems 151, (2017).

70. Cheng, B., et al. Panoptic-DeepLab: A Simple, Strong, and Fast Baseline for Bottom-Up Panoptic Segmentation. arXiv:1911.10194 [cs] (2020).

71. Van der Walt, S. et al. scikit-image: image processing in Python. PeerJ 2, e453 (2014).

72. Pedregosa, F. et al. Scikit-learn: Machine Learning in Python. Journal of Machine Learning Research 12, 2825–2830 (2011).

73. Kim, D. et al. Evaluating the role of Z-stack to improve the morphologic evaluation of urine cytology whole slide images for high-grade urothelial carcinoma: Results and review of a pilot study. Cancer Cytopathology 130, 630–639 (2022).

74. Allison, D. B. et al. Should the BK polyomavirus cytopathic effect be best classified as atypical or benign in urine cytology specimens? Cancer cytopathology 124, 436–442 (2016).

75. Wu, Z. et al. Representing long-range context for graph neural networks with global attention. Advances in Neural Information Processing Systems 34, 13266–13279 (2021).

76. Levy, J. J., Salas, L. A., Christensen, B. C., Sriharan, A. & Vaickus, L. J. PathFlowAI: A High-Throughput Workflow for Preprocessing, Deep Learning and Interpretation in Digital Pathology. Pac Symp Biocomput 25, 403–414 (2020).

77. Sundararajan, M., Taly, A. & Yan, Q. Axiomatic attribution for deep networks. in Proceedings of the 34th International Conference on Machine Learning - Volume 70 3319–3328 (JMLR.org, 2017).

78. Kokhlikyan, N. et al. Captum: A unified and generic model interpretability library for PyTorch. arXiv:2009.07896 [cs, stat] (2020).

79. Falk, T. et al. U-Net: deep learning for cell counting, detection, and morphometry. Nature methods 16, 67–70 (2019).

80. Ronneberger, O., Fischer, P. & Brox, T. U-net: Convolutional networks for biomedical image segmentation. in Medical Image Computing and Computer-Assisted Intervention–MICCAI 2015: 18th International Conference, Munich, Germany, October 5-9, 2015, Proceedings, Part III 18 234–241 (Springer, 2015).

81. He, K., Zhang, X., Ren, S. & Sun, J. Deep Residual Learning for Image Recognition. in 2016 IEEE Conference on Computer Vision and Pattern Recognition (CVPR) 770–778 (2016). doi:10.1109/CVPR.2016.90.

82. Koss, L. G., Deitch, D., Ramanathan, R. & Sherman, A. B. Diagnostic value of cytology of voided urine. Acta cytologica 29, 810–816 (1985).

83. Onur, I., Rosenthal, D. L. & VandenBussche, C. J. Benign-appearing urothelial tissue fragments in noninstrumented voided urine specimens are associated with low rates of urothelial neoplasia. Cancer Cytopathology 123, 180–185 (2015).

84. Thompson, C. G., Kim, R. S., Aloe, A. M. & Becker, B. J. Extracting the variance inflation factor and other multicollinearity diagnostics from typical regression results. Basic and Applied Social Psychology 39, 81–90 (2017).

85. Carvalho, C. M., Polson, N. G. & Scott, J. G. Handling Sparsity via the Horseshoe. In Artificial Intelligence and Statistics 73–80 (PMLR, 2009).

86. McKinley, T. J., Morters, M. & Wood, J. L. N. Bayesian Model Choice in Cumulative Link Ordinal Regression Models. Bayesian Analysis 10, 1–30 (2015).

87. Bender, R. & Grouven, U. Ordinal Logistic Regression in Medical Research. J R Coll Physicians Lond 31, 546–551 (1997).

88. OpenSeadragon. http://openseadragon.github.io/.

89. Wolfson, W. L. & Rosenthal, D. L. Cell clusters in urinary cytology. Acta Cytol 22, 138–141 (1978).

90. Mikou, P. et al. Evaluation of the Paris System in atypical urinary cytology. Cytopathology 29, 545–549 (2018).

91. Kurtycz, D. F. et al. Paris interobserver reproducibility study (PIRST). Journal of the American Society of Cytopathology 7, 174–184 (2018).

92. Kurtycz, D. F. I., Sundling, K. E. & Barkan, G. A. The Paris system of Reporting Urinary Cytology: Strengths and opportunities. Diagnostic Cytopathology 48, 890–895 (2020).

93. Bakkar, R. et al. Impact of the Paris system for reporting urine cytopathology on predictive values of the equivocal diagnostic categories and interobserver agreement. Cytojournal 16, (2019).

94. Hassan, M. et al. Impact of Implementing the Paris System for Reporting Urine Cytology in the Performance of Urine Cytology: A Correlative Study of 124 Cases. American Journal of Clinical Pathology 146, 384–390 (2016).

95. Pantanowitz, L. Automated pap tests. Practical Informatics for Cytopathology 147–155 (2014).

96. Yamashita, S. et al. Urethral recurrence following neobladder in bladder cancer patients. The Tohoku Journal of Experimental Medicine 199, 197–203 (2003).

97. Pierconti, F. et al. DNA methylation analysis in urinary samples: A useful method to predict the risk of neoplastic recurrence in patients with urothelial carcinoma of the bladder in the high-risk group. Cancer Cytopathology n/a,.

98. Shalata, A. T. et al. Predicting Recurrence of Non-Muscle-Invasive Bladder Cancer: Current Techniques and Future Trends. Cancers 14, 5019 (2022).

99. Soorojebally, Y. et al. Urinary biomarkers for bladder cancer diagnosis and NMIBC follow-up: a systematic review. World J Urol 41, 345–359 (2023).

100. Lujan, G. et al. Dissecting the business case for adoption and implementation of digital pathology: a white paper from the digital pathology association. Journal of Pathology Informatics 12, 17 (2021).

101. Acs, B. & Rimm, D. L. Not just digital pathology, intelligent digital pathology. JAMA oncology 4, 403–404 (2018).

102. Jones-Hall, Y. Digital pathology in academia: Implementation and impact. Lab Animal 50, 229–231 (2021).

103. Olswang, L. B. & Prelock, P. A. Bridging the gap between research and practice: Implementation science. Journal of Speech, Language, and Hearing Research 58, S1818– S1826 (2015).

104. Li, R. C., Asch, S. M. & Shah, N. H. Developing a delivery science for artificial intelligence in healthcare. *npj Digit*. Med. 3, 1–3 (2020).

105. Char, D. S., Abràmoff, M. D. & Feudtner, C. Identifying ethical considerations for machine learning healthcare applications. The American Journal of Bioethics 20, 7–17 (2020).

106. Jackson, B. R., et al. The Ethics of Artificial Intelligence in Pathology and Laboratory Medicine: Principles and Practice. Acad Pathol 8, (2021).

107. Chauhan, C. & Gullapalli, R. R. Ethics of AI in pathology: current paradigms and emerging issues. The American journal of pathology 191, 1673–1683 (2021).

108. Baxi, V., Edwards, R., Montalto, M. & Saha, S. Digital pathology and artificial intelligence in translational medicine and clinical practice. Modern Pathology 35, 23–32 (2022).

109. Niazi, M. K. K., Parwani, A. V. & Gurcan, M. N. Digital pathology and artificial intelligence. The Lancet Oncology 20, e253–e261 (2019).

110. Bouyssoux, A., Fezzani, R. & Olivo-Marin, J.-C. Cell Instance Segmentation Using Z-Stacks in Digital Cytology. in 2022 IEEE 19th International Symposium on Biomedical Imaging (ISBI) 1–4 (2022). doi:10.1109/ISBI52829.2022.9761495.

111. Vaickus, L. J. & Tambouret, R. H. Young investigator challenge: The accuracy of the nuclear-to-cytoplasmic ratio estimation among trained morphologists. Cancer Cytopathol 123, 524–530 (2015).

112. Butke, J. et al. End-to-end Multiple Instance Learning for Whole-Slide Cytopathology of Urothelial Carcinoma. in Proceedings of the MICCAI Workshop on Computational Pathology 57–68 (PMLR, 2021).

113. Lu, M. Y. et al. Data-efficient and weakly supervised computational pathology on whole-slide images. Nature Biomedical Engineering 1–16 (2021) doi:10.1038/s41551-020-00682-w.

114. Vargas, V. M., Gutiérrez, P. A. & Hervás-Martínez, C. Cumulative link models for deep ordinal classification. Neurocomputing 401, 48–58 (2020).

